# Cardi-Ankle Vascular Index Optimizes Ischemic Heart disease Diagnosis

**DOI:** 10.1101/2024.07.03.24309877

**Authors:** Basheer Abdullah Marzoog, Daria Gognieva, Peter Chomakhidze, Philipp Kopylov

## Abstract

**Background:** Ischemic heart disease (IHD) has the highest mortality rate in the globe in between the other cardiovascular diseases (CVD). This returns to the poor diagnostic and therapeutic strategies including the primary prevention techniques.

**Aims:** To assess the changes in the cardio-ankle vascular index (CAVI) in patients with vs without IHD confirmed by stress computed tomography myocardial perfusion (CTP) imaging with vasodilatation stress-test (Adenosine triphosphate).

**Objectives:** IHD often has preventable risk factors and causes that lead to the appearance of the disease. However, the lack of appropriate diagnostic and prevention tools remains a global challenge in or era despite current scientific advances.

**Material and methods:** A single center observational study included 80 participants from Moscow. The participants aged ≥ 40 years and given a written consent to participate in the study. Both groups, G1=31 with vs. G2 = 49 without post stress induced myocardial perfusion defect, received cardiologist’s consultation, anthropometric measurements, blood pressure and pulse rate, echocardiography, CAVI and performing bicycle ergometry. For statistical analysis, descriptive statistics, t-test independent by groups and dependent by numerical variables for repeated analysis for the same patients, Pearson’s correlation coefficient, multivariate ANOVA test, and for clarification purposes, diagrams and bar figures were used. For performing the statistical analysis, used the Statistica 12 programme (StatSoft, Inc. (2014). STATISTICA (data analysis software system), version 12. www.statsoft.com.) and the IBM SPSS Statistics, version 28.0.1.1 (14).

**Results:** The mean age of the participants 56.28, standard deviation (Std.Dev. ± 10.601). Mean CAVI in the IHD group 8.509677 (Std.Dev. ± 0.975057208) vs 7.994898 (Std.Dev. ± 1.48990509) in the non-IHD group. The mean estimated biological age of the arteries according to the results of the CAVI in the first group 61.2258 years vs 53.5102 years in the second group. The Mean brachial-ankle pulse (Tba) in the IHD group 82.0968 vs 89.0102 in the second group. The mean heart-ankle pulse wave velocity (haPWV; m/s) in the IHD group was 0.9533 vs 0.8860 in the second group. Regression analysis demonstrated that the dependent variable, the CAVI parameter, have no significant effect on the development of stress-induced myocardial perfusion defect, regression coefficient 95.316, p>0.05. The CAVI showed 64 % diagnostic accuracy for the IHD.

**Conclusion:** The CAVI parameter showed no statistical difference between the participants with IHD vs without. The CAVI parameter can be used as an axillary method for improving the diagnosis of IHD.

**Other:** Additional indicators associated with IHD include the Tba and haPWV parameters, higher in patients with IHD.

**Graphical abstract:** 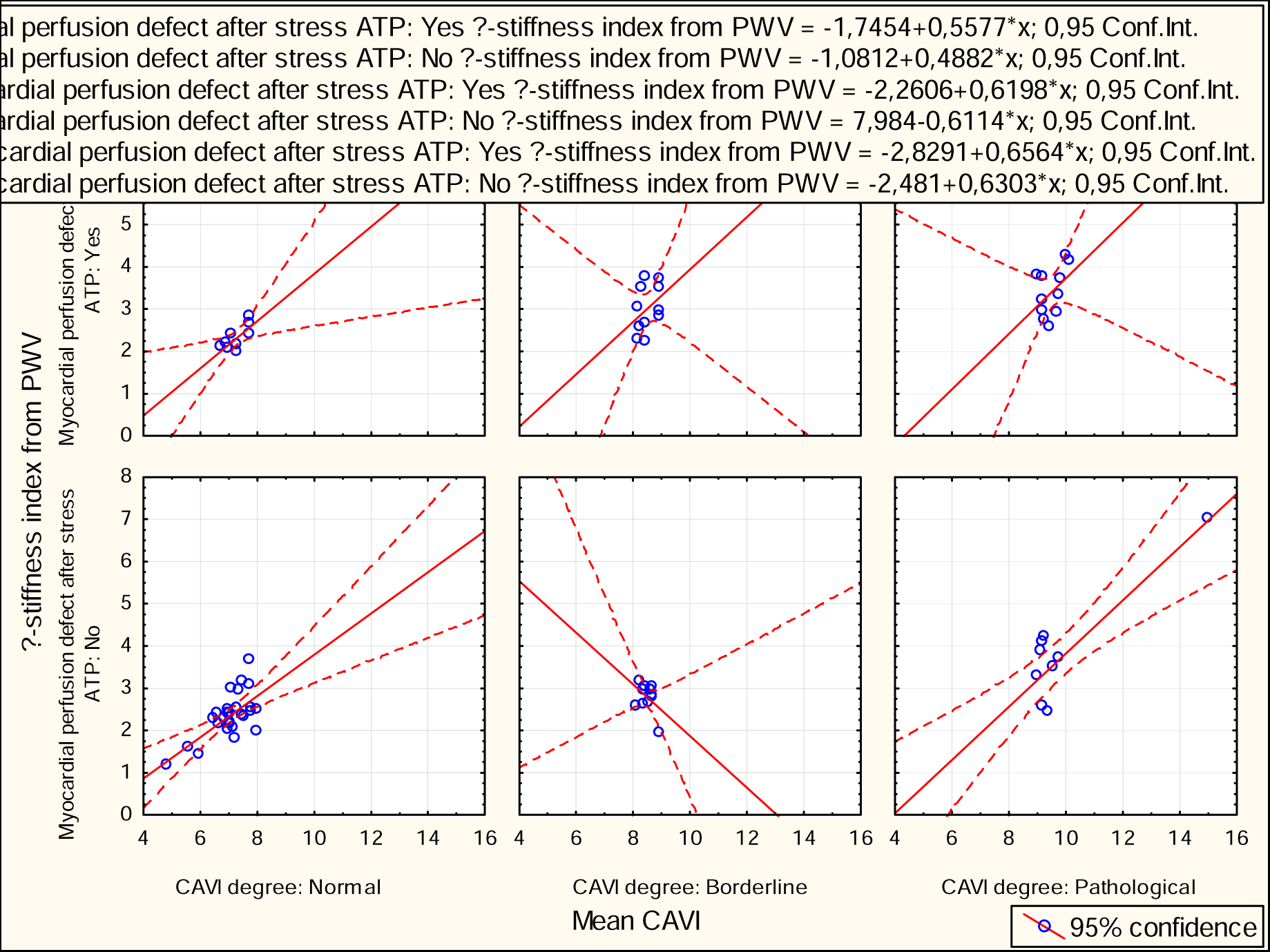

## Introduction

Ischemic heart disease remains the main challenge in terms of mortality and morbidity despite advances in the used methods for diagnosis and prevention. However, the early prevention in terms of evaluation of ischemic heart disease in early period was still underestimated. The current attention of scientists paid to the prevention rather than diagnosis and treatment. In this manner, the scientific community developed several cost-effective methods to be confirmed for the clinical use for early prevention of ischemic heart disease, including the use of single-channel electrocardiography and exhaled breath analysis in coronary heart disease prevention [1].

Vectorcardiography, a technique that records cardiac electrical activity as closed loops, can be useful for training in electrocardiography and detecting cardiac ischemia [2,3]. Portable and fast electrode placement devices allow for good-quality ECG tracings, making single-channel ECG accessible and efficient [2].

The usage of CAVI in ischemic heart disease has a challenging role and requires further investigation and elucidation.

## Material and methods

A prospective single center cohort study included 80 participants. According to the results of the CTP, the participates divided in to two groups. The first group of participants with stress-induced myocardial perfusion defect (n=31) and the second group without stress induced myocardial perfusion defect (n = 49) in the CTP. Participants are randomly chosen. Written consent has been taken from the participants. The study registered on clinicaltrials.gov (NCT06181799), and the study approved by the ethical commitment of the Sechenov University, Russia, from “Ethics Committee Requirement № 19-23 from 26.10.2023”. The study evaluated continuous and categorical variables. The continuous variables included; age, pulse at rest, systolic blood pressure (SBP) at rest, diastolic blood pressure (DBP) at rest, body weight, height, maximum heart rate (HR) on physical stress test, watt (WT) on physical stress test, metabolic equivalent (METs) on physical stress test, reached percent on physical stress test, ejection fraction (EF %) on echocardiography, estimated vessel age, right cardio-ankle vascular index (R-CAVI), left Cardio-ankle vascular index (L-CAVI), mean CAVI (=(right-CAVI + left-CAVI)/2), right ankle-brachial index (RABI), left ankle-brachial index (LABI), mean ankle-brachial index (ABI), mean SBP brachial (SBPB) (=(right SBPB+ left SBPB)/2), mean DBPB (=(right DBPB + left DBPB)/2), BP right brachial (BPRB) (=(SBP+DBP)/2), BP left brachial (BPLB) (=(SBP+DBP)/2), mean BPB (=(BPRB+ BPLB)/2), BP right ankle (BPRA) (=(SBP+DBP)/2), BP left ankle (BPLA) (=(SBP+DBP)/2), mean BPA (=(BPRA+ BPLA) /2), right brachial pulse (RTb), left brachial pulse (LTb), mean Tb (=(LTb+ RTb)/2), right brachial-ankle pulse (Tba), left brachial-ankle pulse (Tba), mean Tba (= (left Tba+right Tba)/2), length heart-ankle (Lha in cm), heart-ankle pulse wave velocity (haPWV = Lha/(mean left Tba+ mean right Tba); m/s), β-stiffness index from PWV (=2*1050*(haPWV)
^2*LN((mean SBPB *133,32/ mean DBPB *133,32))/((mean SBPB*133,32)-(mean DBPB *133,32)), creatinine (µmol/L), and eGFR (2021 CKD-EPI Creatinine). Categorical variables included; gender, obesity stage, smoking, concomitant disease, coronary artery, hemodynamically significant (>60%), myocardial perfusion defect after stress ATP, myocardial perfusion defect before stress ATP, atherosclerosis in other arteries (Yes/No), carotid atherosclerosis, brachiocephalic atherosclerosis, arterial hypertension (AH), stage of the AH, degree of the AH, risk of cardiovascular disease (CVD), stable coronary artery disease (SCAD), functional class (FC) by Watt and by METs, reaction type to stress test(positive/negative), reason of discontinuation of the stress test, CAVI degree, and ABI degree.

## The selection criteria involved

The inclusion criteria;

1. Participants age ≥ 40 years.
2. Participants with intact mental and physical activity.
3. Written consent to participate in the study, take blood samples, and anonymously publish the results of the study.
4. The participants in the control group are individuals without coronary artery disease, confirmed by the absence of the stress-induced myocardial perfusion defect on adenosine triphosphate stress myocardial perfusion computed tomography ((by using contrast-enhanced multislice spiral computed tomography (CE-MSCT) using adenosine triphosphate (ATP)).
5. Participants in the experimental group are individuals with coronary artery disease, confirmed by stress-induced myocardial perfusion defect on the adenosine triphosphate stress myocardial perfusion computed tomography.

Exclusion criteria:

1. Failure of the stress test for reasons unrelated to heart disease
2. Reluctance to continue participating in the study.

Non-inclusion criteria

1. Pregnancy.
2. Diabetes mellitus
3. Presence of signs of acute coronary syndrome (myocardial infarction in the last two days), history of myocardial infarction;
4. Active infectious and non-infectious inflammatory diseases in the exacerbation phase;
5. Respiratory diseases (bronchial asthma, chronic bronchitis, cystic fibrosis);
6. Acute thromboembolism of pulmonary artery branches;
7. Aortic dissection;
8. Critical anatomical heart defects;
9. Active oncopathology;
10. Decompensation phase of acute heart failure;
11. Neurological pathology (Parkinson’s disease, multiple sclerosis, acute psychosis, Guillain-Barré syndrome);
12. Cardiac arrhythmias that do not allow exercise ECG testing (Wolff-Parkinson-White syndrome, Sick sinus syndrome, AV block of II-III-degree, persistent ventricular tachycardia);
13. Diseases of the musculoskeletal system that prevent passing a stress test (bicycle ergometry);
14. Allergic reaction to iodine and/or adenosine triphosphate.

## Data collection

Both groups pass a vessel stiffness test and pulse wave recording as well as vascular age using Fukuda Denshi device (VaSera VS-1500; Japan). Cuffs placed to assess vascular stiffness (CAVI parameter) and vascular age as well as the ancle-brachial index [6]. Measurements performed in a quiet room at a stable temperature 24–26 °C; exercise, smoking, discontinue chronotropic medications (beta-blockers and non-dihydropyridine drugs; verapamil, diltiazem) and food avoided 2–3 hours prior to the measurement [7].

Subsequently, participants pass the exercise bicycle ergometry test (SCHILLER CS200 device; Bruce protocol or modified Bruce protocol). According to the metabolic equivalent; Mets-ВT (ВТ), the functional class (FC) in participants with determined positive stress test results determined. During the bicycle ergometry, the participants monitored with 12-lead ECG and manual blood pressure measurement, 1 time each 2 minutes. The rest time ECG and blood pressure monitoring continue for at least five minutes after the end of the stress bicycle ergometry test.

The procedure is stopped if an increase in systolic blood pressure <220 mmHg, horizontal or down on the ECG ≥ 1 mm, typical heart pain during test, ventricular tachycardia or atrial fibrillation, or other significant heart rhythm disorders were found. Furthermore, stop the procedure if the target heart rate (86% of the 220-age) is reached.

Before performing CTP, all the participants present results of the venous creatinine level, eGFR (estimated glomerular filtration rate) according to the 2021 CKD-EPI Creatinine > 30 ml/min/1,73 m2, according to the recommendation for using this formula by the National kidney foundation and the American Society of Nephrology [8–11].

Participants from both groups got catheterization in the basilar vein or the radial vein for injection of contrast and ATP to perform a pharmacological stress test in the heart by increasing heart rate during the computer tomography imaging of stressed myocardial perfusion.

Computer tomography was performed on Canon scanner with 640 slice, 0,5 mm thickness of slice, with contrast (Omnipaque, 50 ml), injected two times: in rest to get images for myocardial perfusion before test, and in 20 mints just after ATP had been injected in dose according to body weight.

The results of myocardial perfusion considered positive if there was a perfusion defect after the stress test or worsen the already existing perfusion defect in the rest phase.

Statistical processing was carried out using the Statistica 12 programme. (StatSoft, Inc. (2014). STATISTICA (data analysis software system), version 12. www.statsoft.com.).

## Statistical Analysis

For quantitative parameters, the nature of the distribution (using the Shapiro-Wilk test), the mean, the standard deviation, the interquartile, the 95% confidence interval, the minimum and maximum values were determined. For categorical and qualitative features, the proportion and absolute number of values were determined. Comparative analysis for normally distributed quantitative traits was carried out on the basis of Welch’s t-test (2 groups); for abnormally distributed quantitative traits, using the Mann-Whitney U-test (2 groups). Comparative analysis of categorical and qualitative features was carried out using the Pearson X-square criterion, in case of its inapplicability, using the exact Fisher test. For the risk of CAVI on the development of stress-induced myocardial perfusion defect, we used multivariate regression analysis using the SPSS programme. For assessing the diagnostic accuracy of the CAVI parameter for stress-induced myocardial perfusion defect, we used the ROC and it is AUC analysis. P considered significant at <0.05.

## Results

The descriptive characteristics of the sample were shown as both groups and then each group separately in tables for a full representation of the results. The characteristics of the continuous variables of the sample described in the table below. (*Table 1)*

**Table 1:**
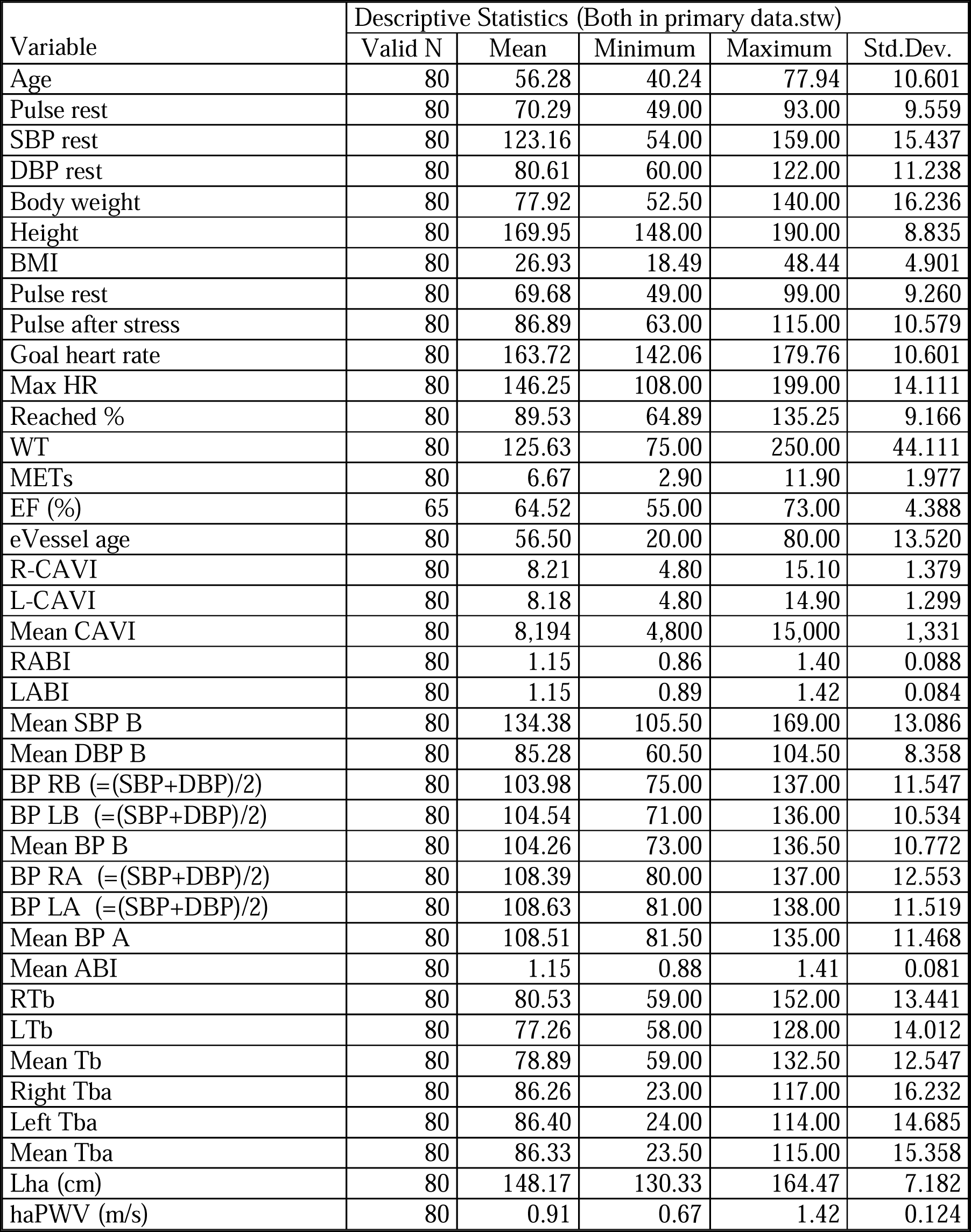
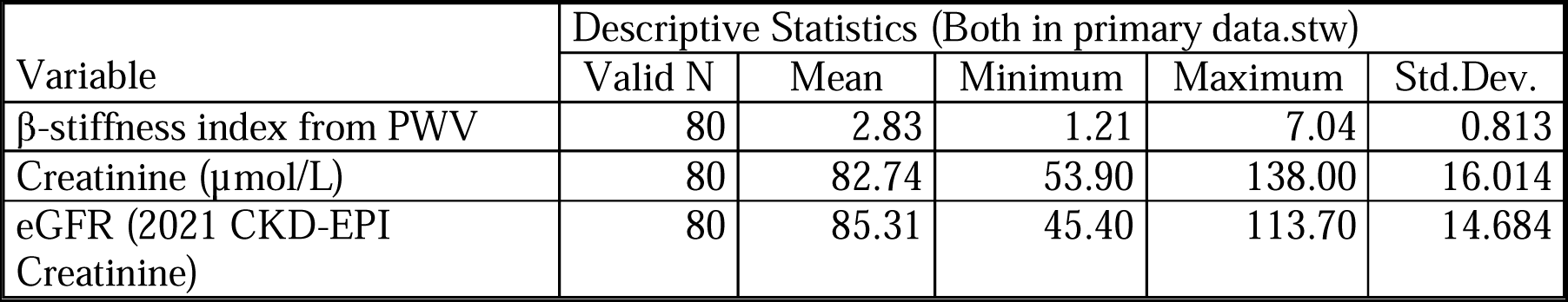
The features of the continues variables of the sample represented in the table.

The comparative characteristics of the sample represented in the below tables based on the presence or absence of the stress induced myocardial perfusion defect of the CTP imaging with the adenosine triphosphate. (*Table 2A-B*)

**Table 2A:**
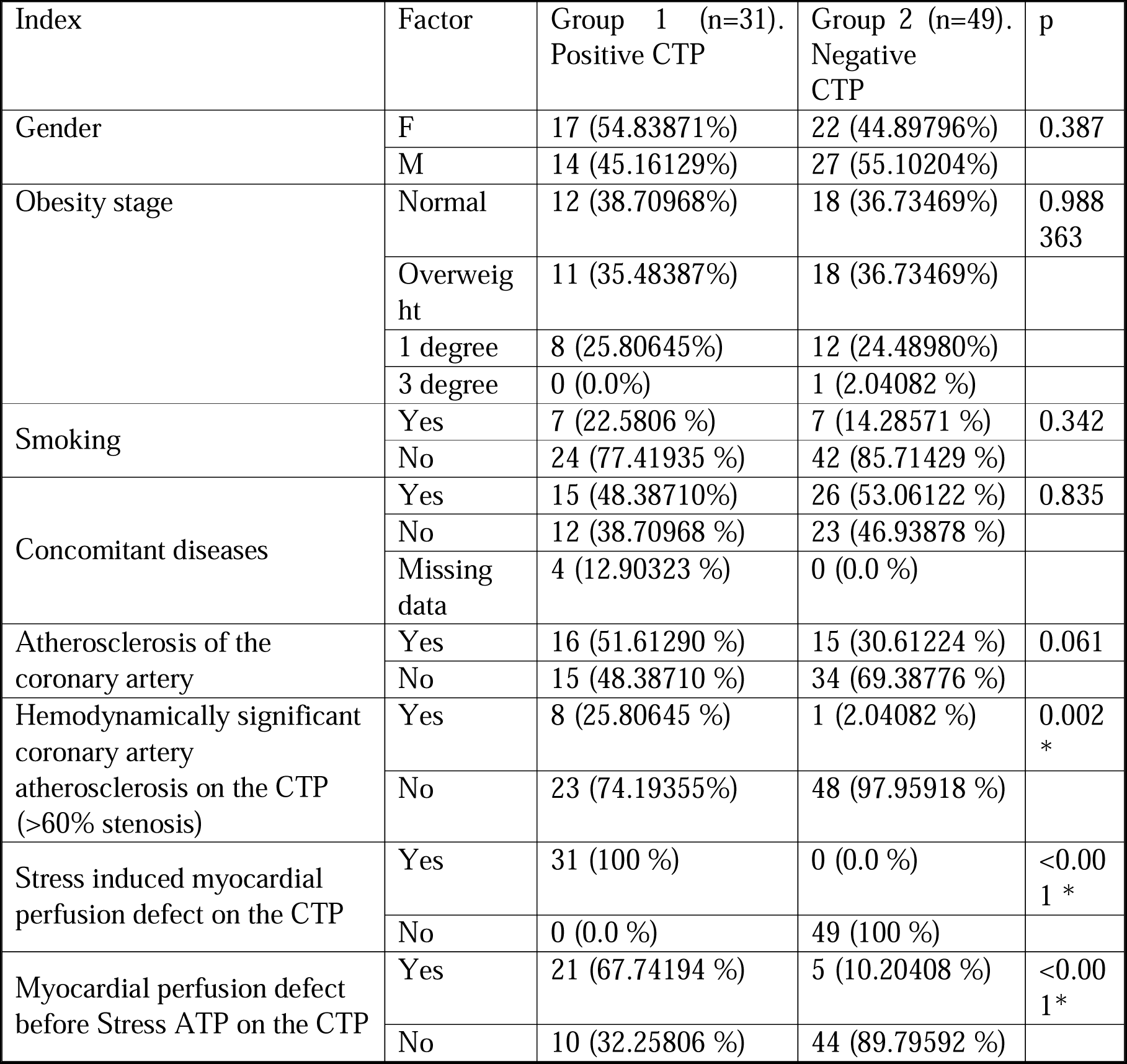
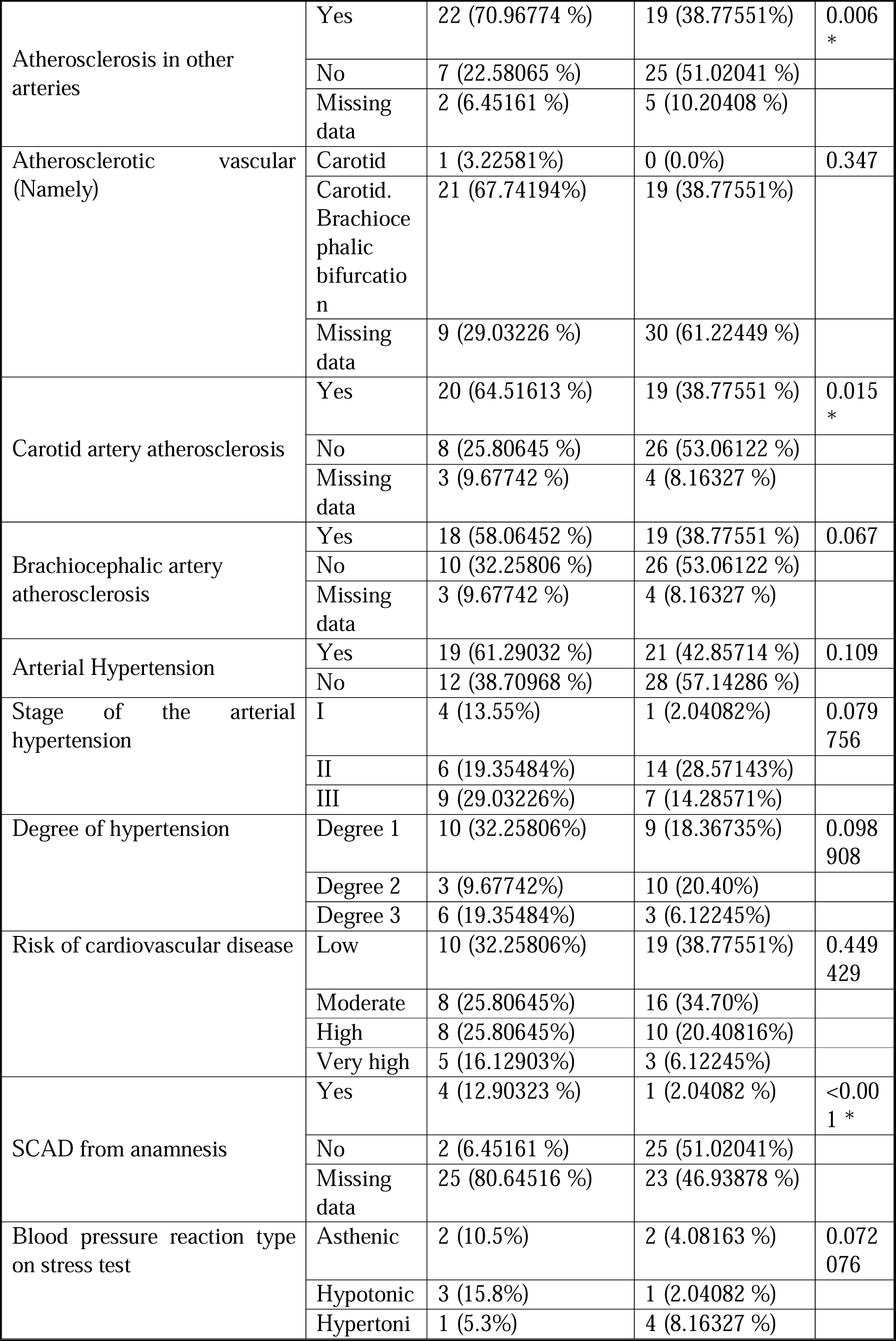
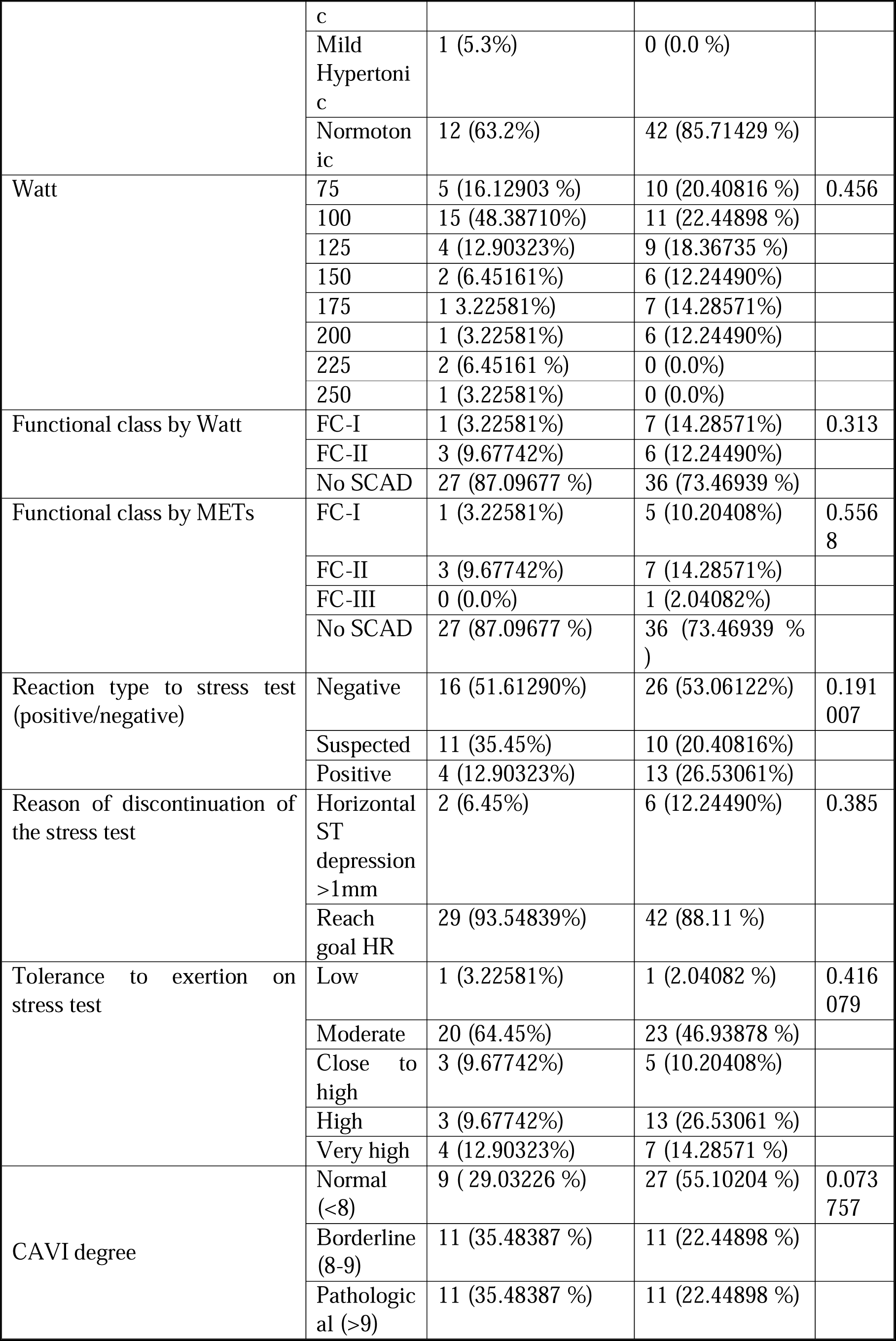
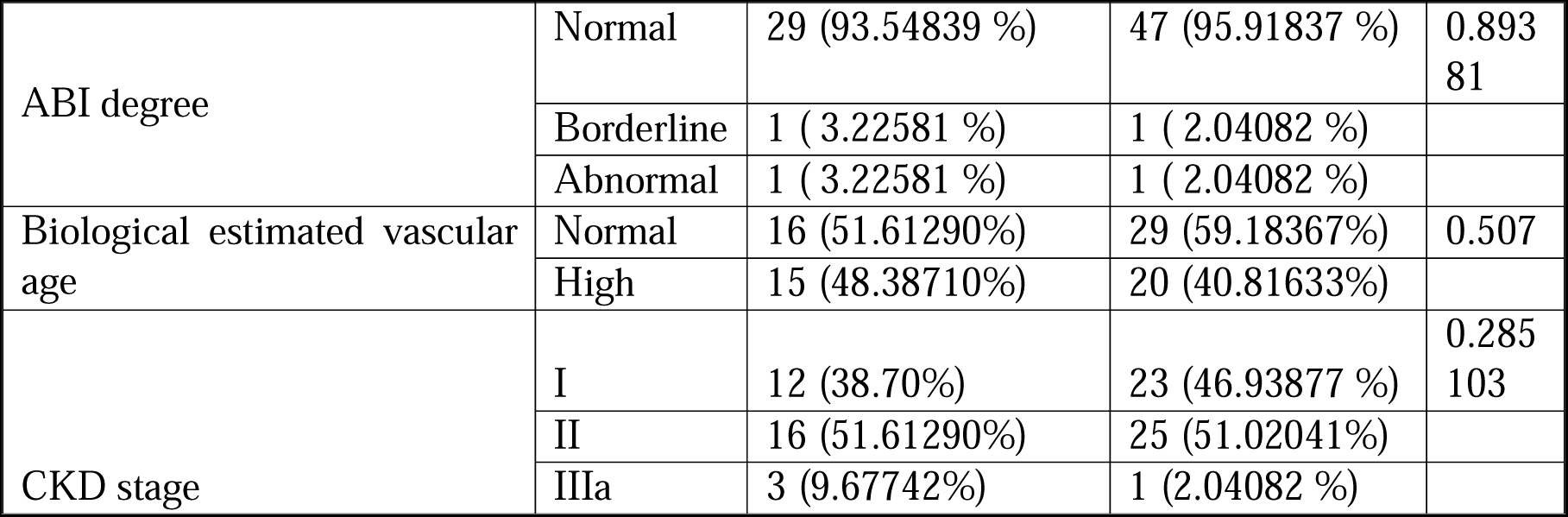
Categorical variables presented in absolute and relative study values for the true incidence of the stated factor. X^2^ test used for the analysis of categorical variables. * Values of statically significant differences. Abbreviations METs; metabolic equivalent, CPT; stress myocardial perfusion computer tomography imaging.

**Table 2B:**
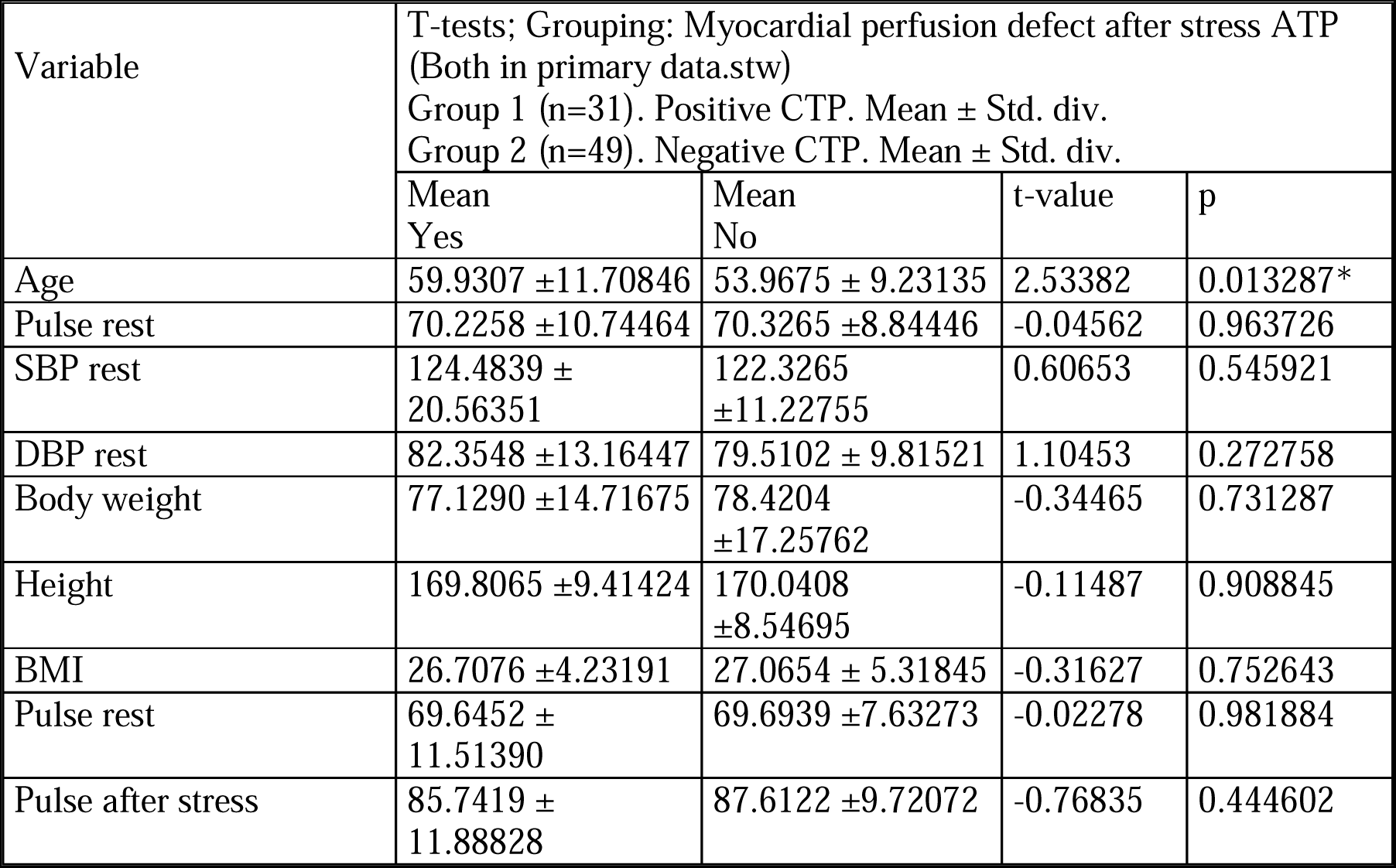
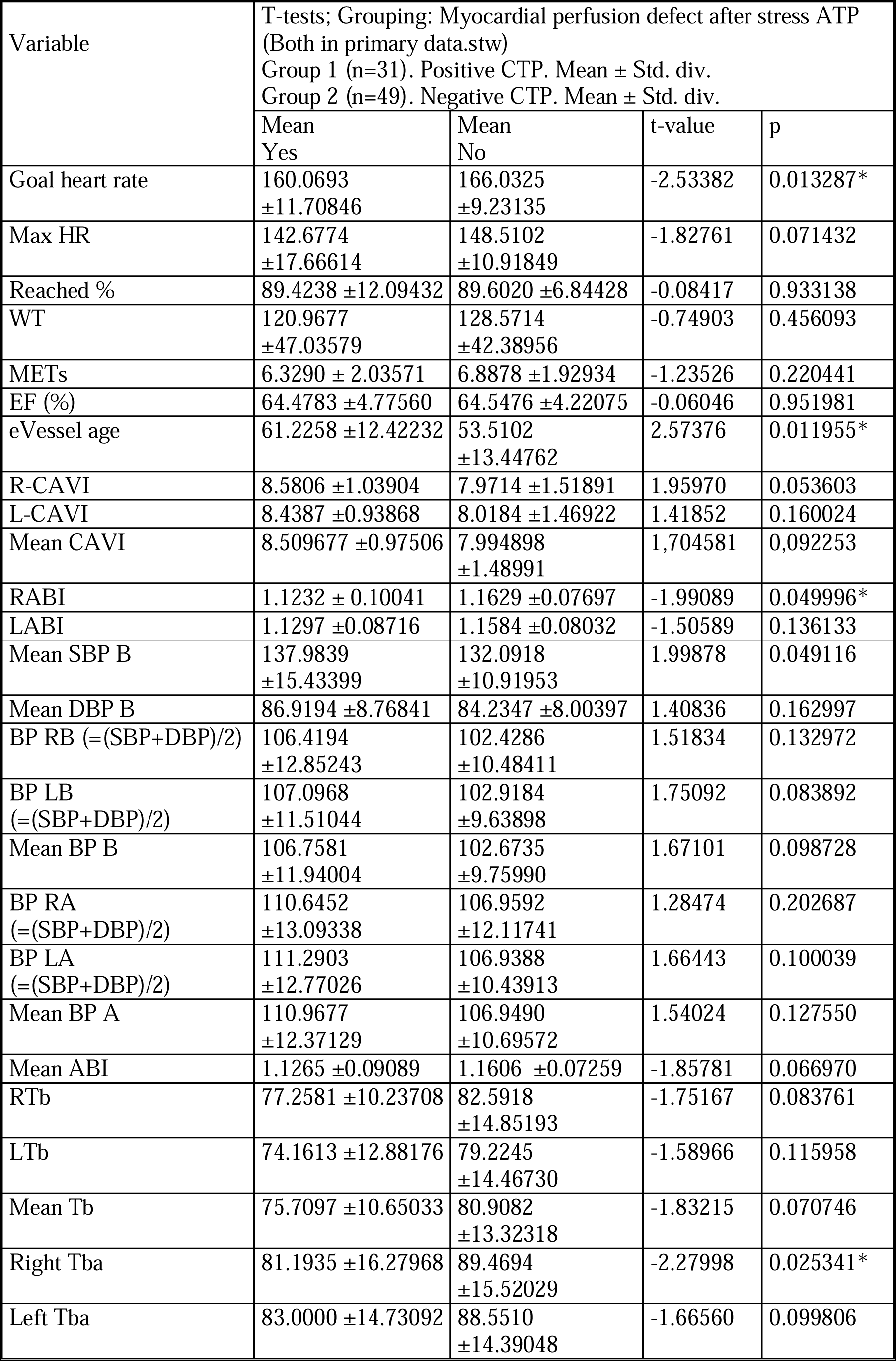
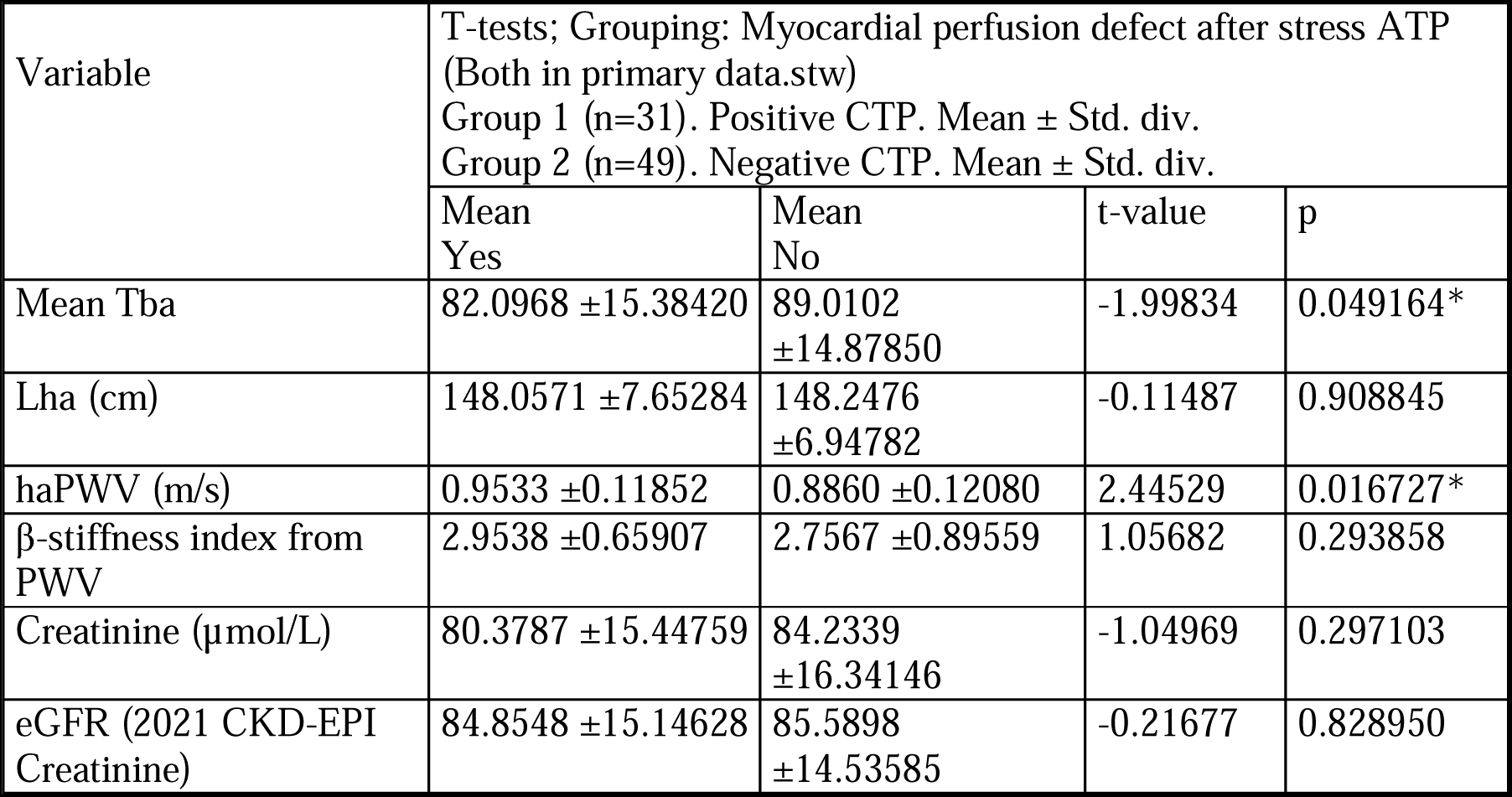
Continuous variables of the sample presented as mean ± standard deviation (Std. div.), Student test as independent variables used. * Values of statically significant differences. Abbreviations: SBP; systolic blood pressure, DBP; diastolic blood pressure, BMI, body mass index, HR; heart rate, METs; metabolic equivalent, R-CAVI; right cardio-ankle vascular index, L-CAVI; left Cardio-ankle vascular index, RABI; right ankle-brachial index, LABI; left ankle-brachial index, SBP B; systolic blood pressure brachial, DBP B; diastolic blood pressure brachial, BP RB; blood pressure right brachial, BP RA; blood pressure right ankle, BP LA; blood pressure left ankle, BP A; blood pressure ankle, ABI; ankle-brachial index, RTb; right brachial pulse, LTb; left brachial pulse, Tb; mean brachial pulse, Tba; mean brachial-ankle pulse, Lha (cm); length heart-ankle, haPWV (m/s); heart-ankle pulse wave velocity.

The bivariant continuous variables of the sample are represented in the following tables. (*Table 3A-3 3M*)

**Table 3:**
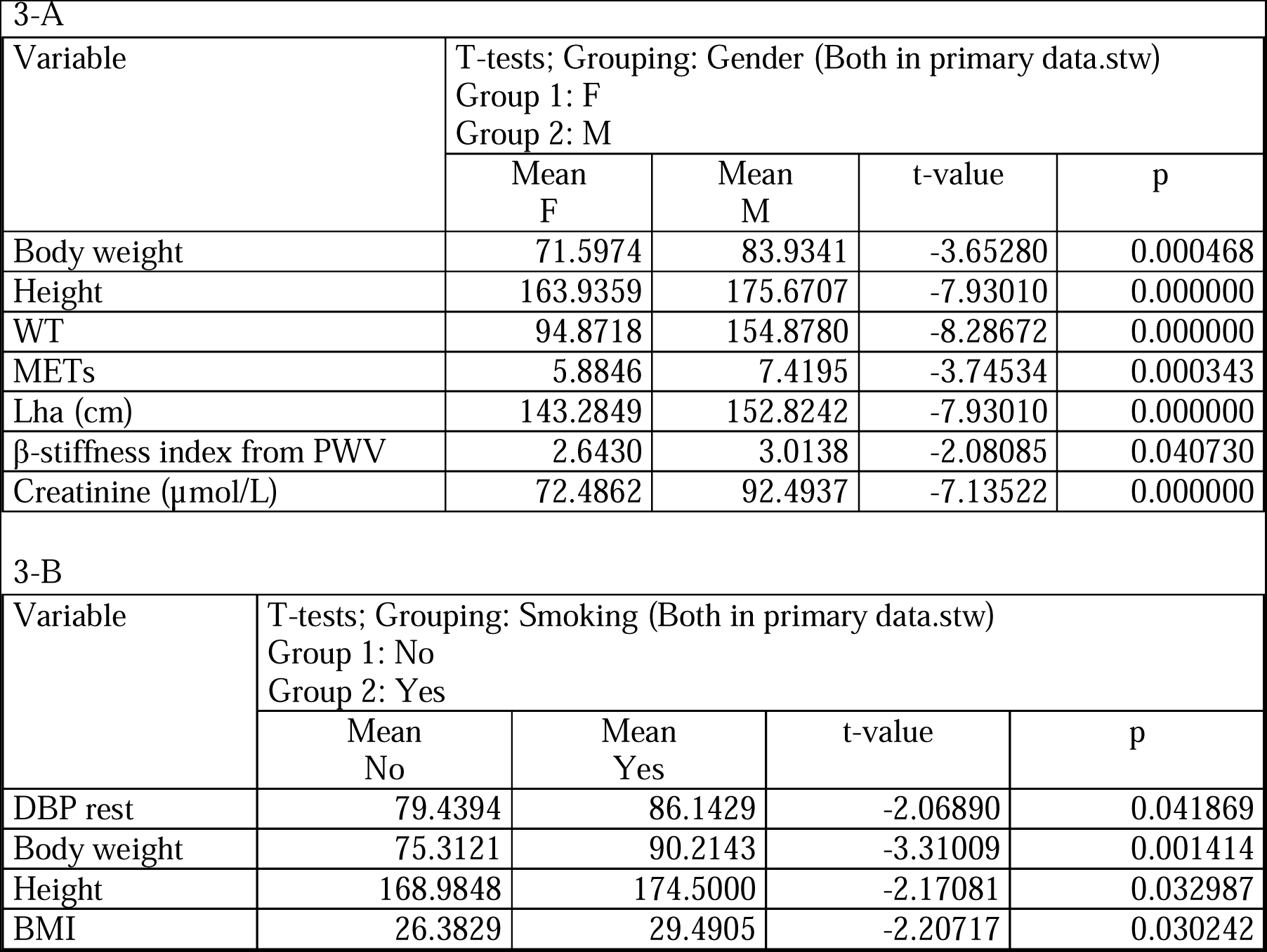
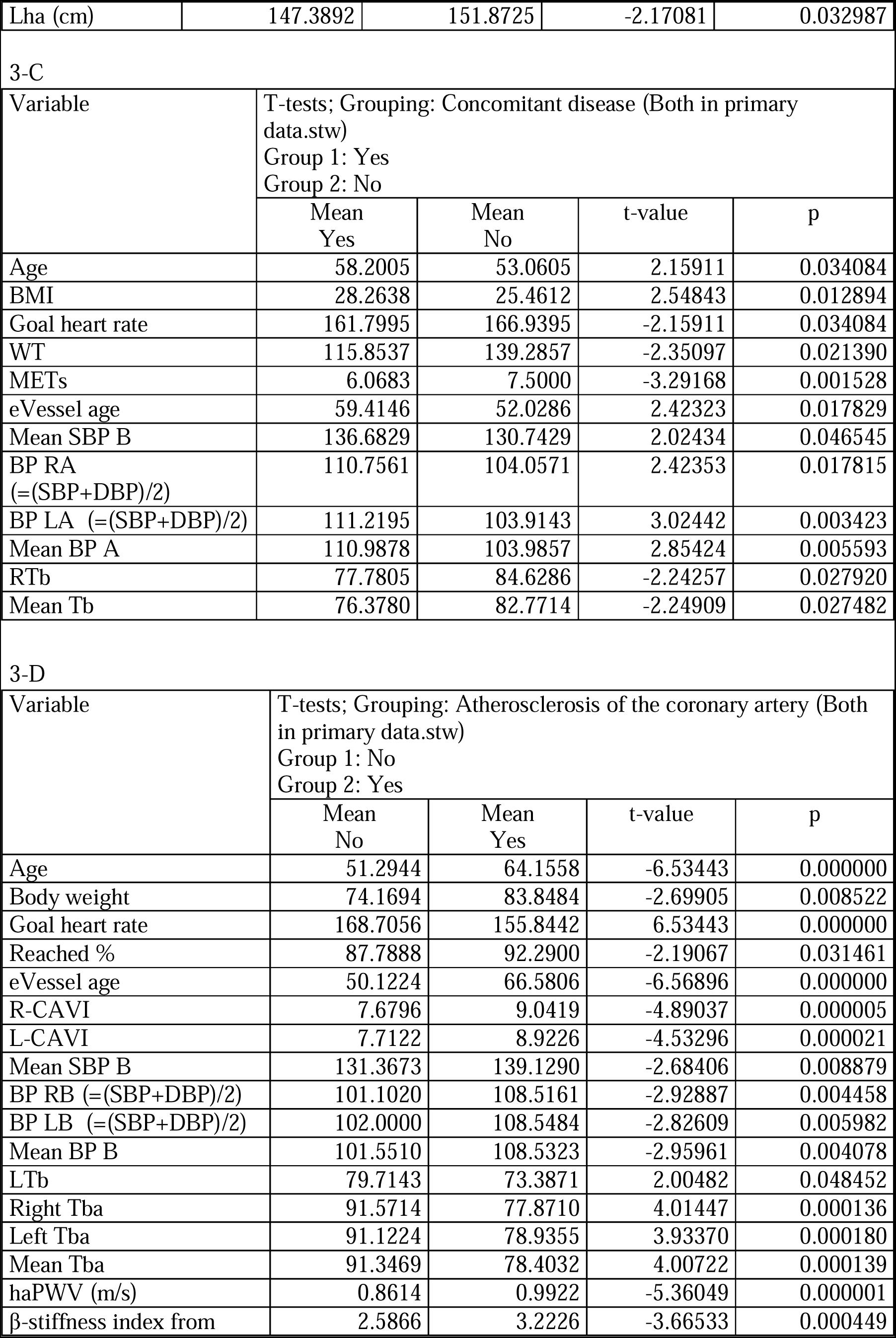
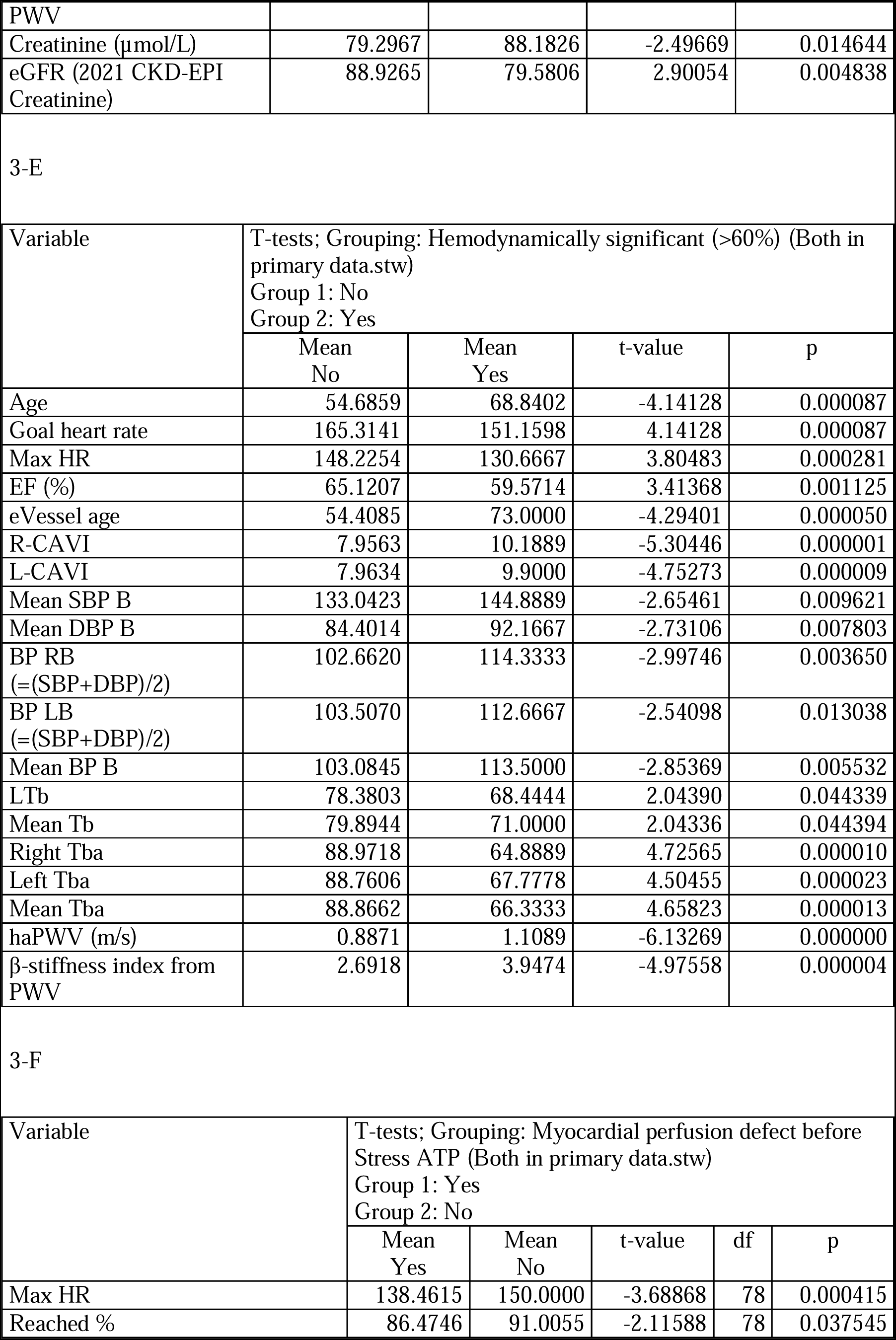
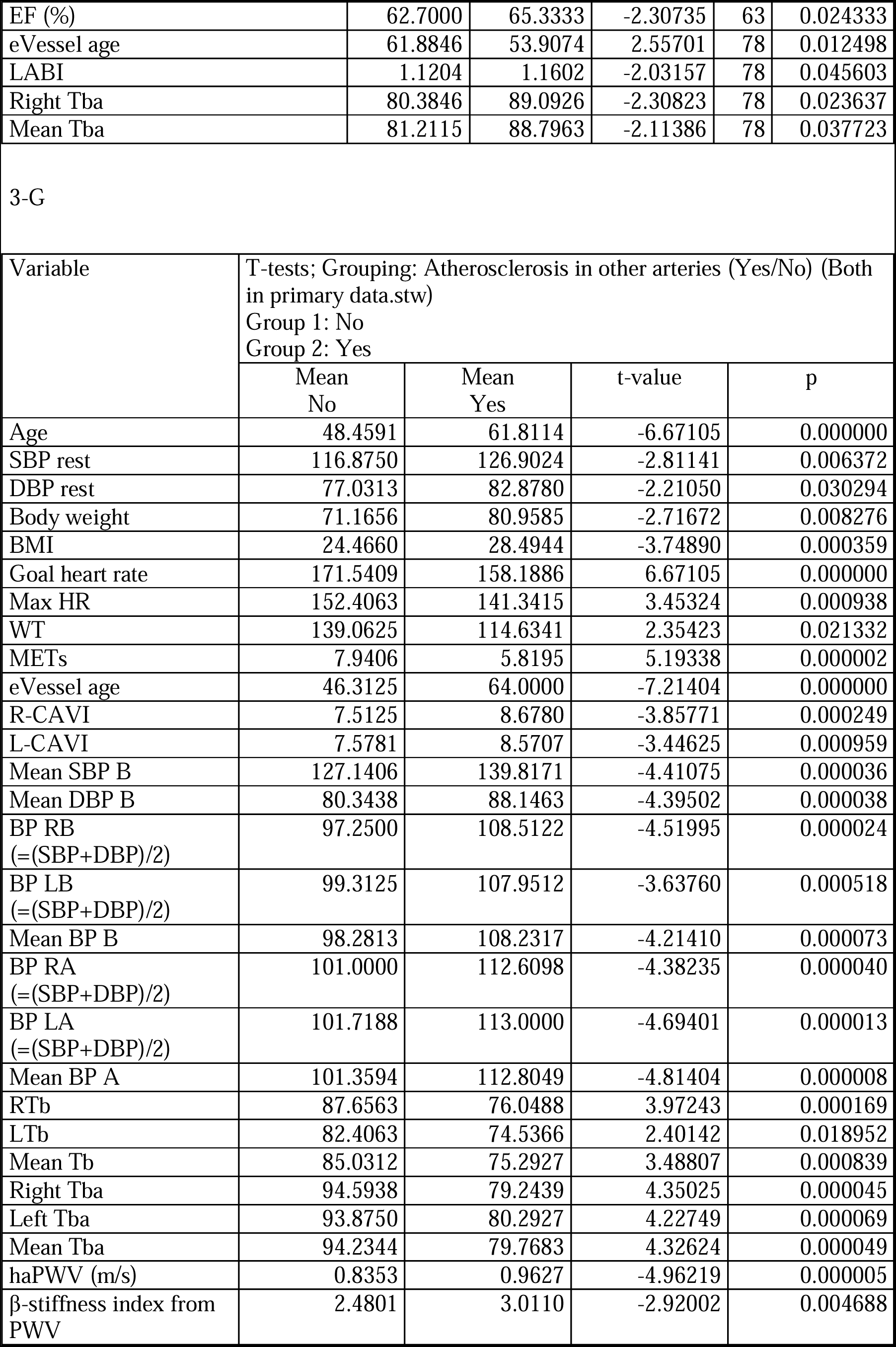
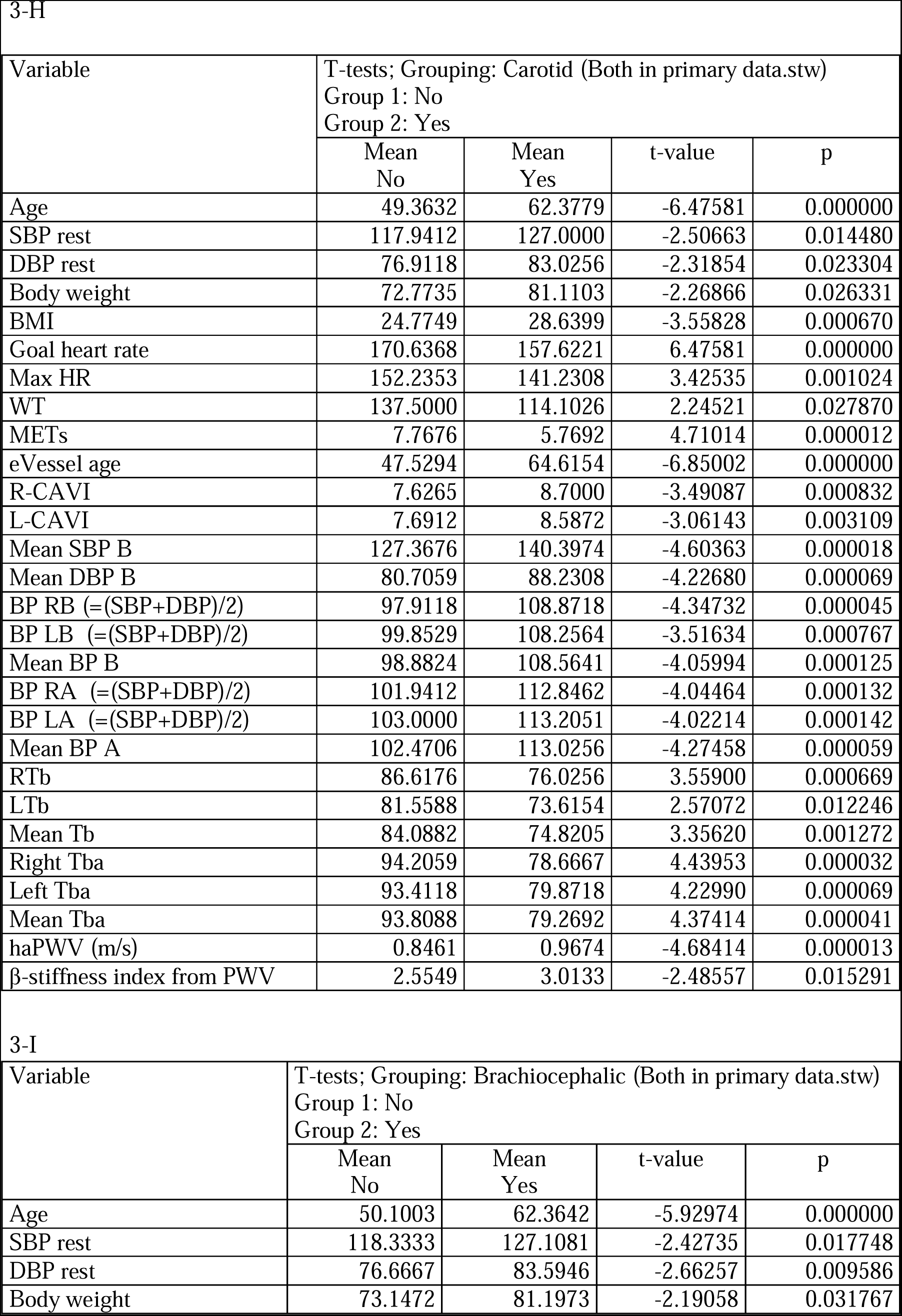
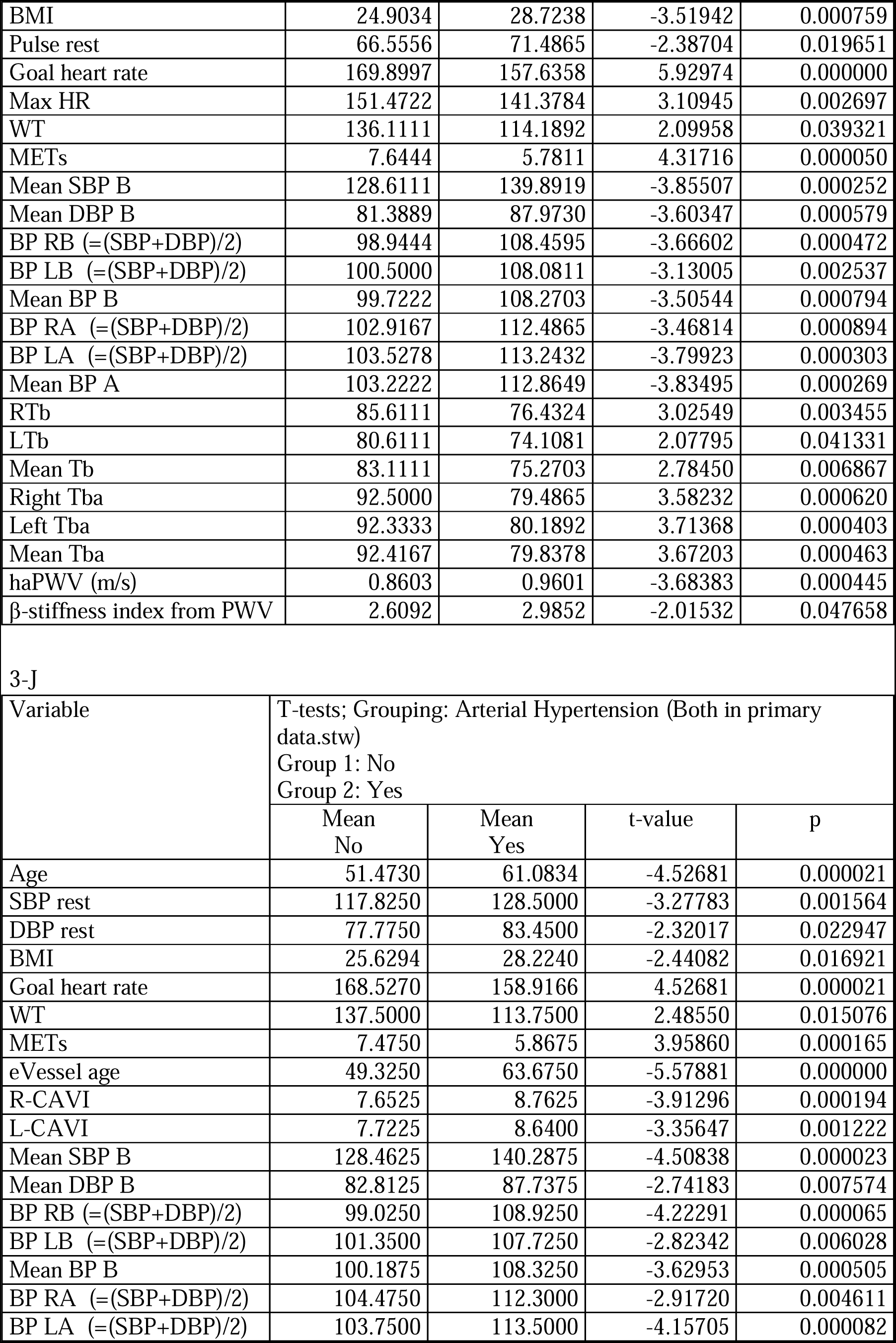
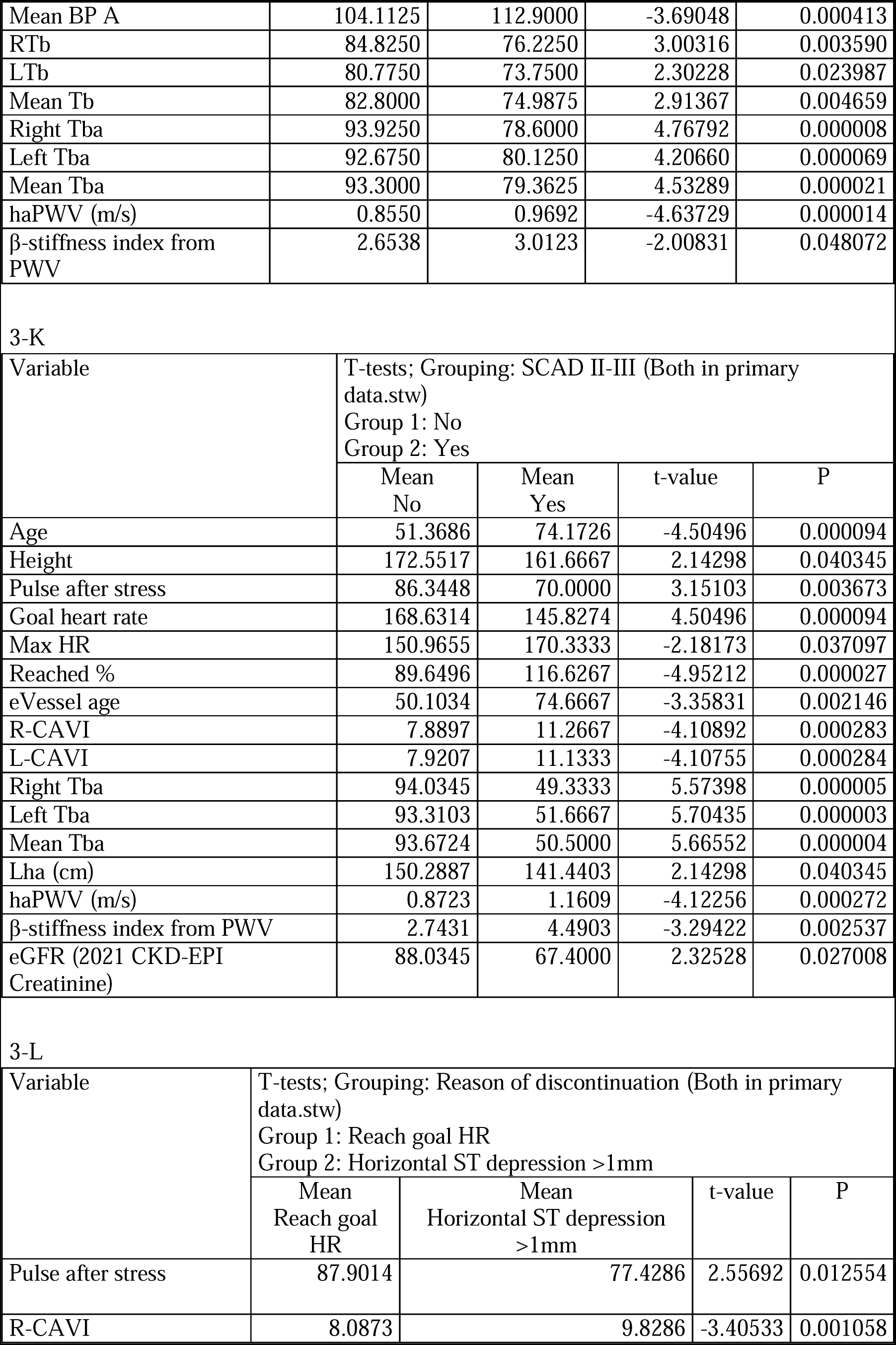
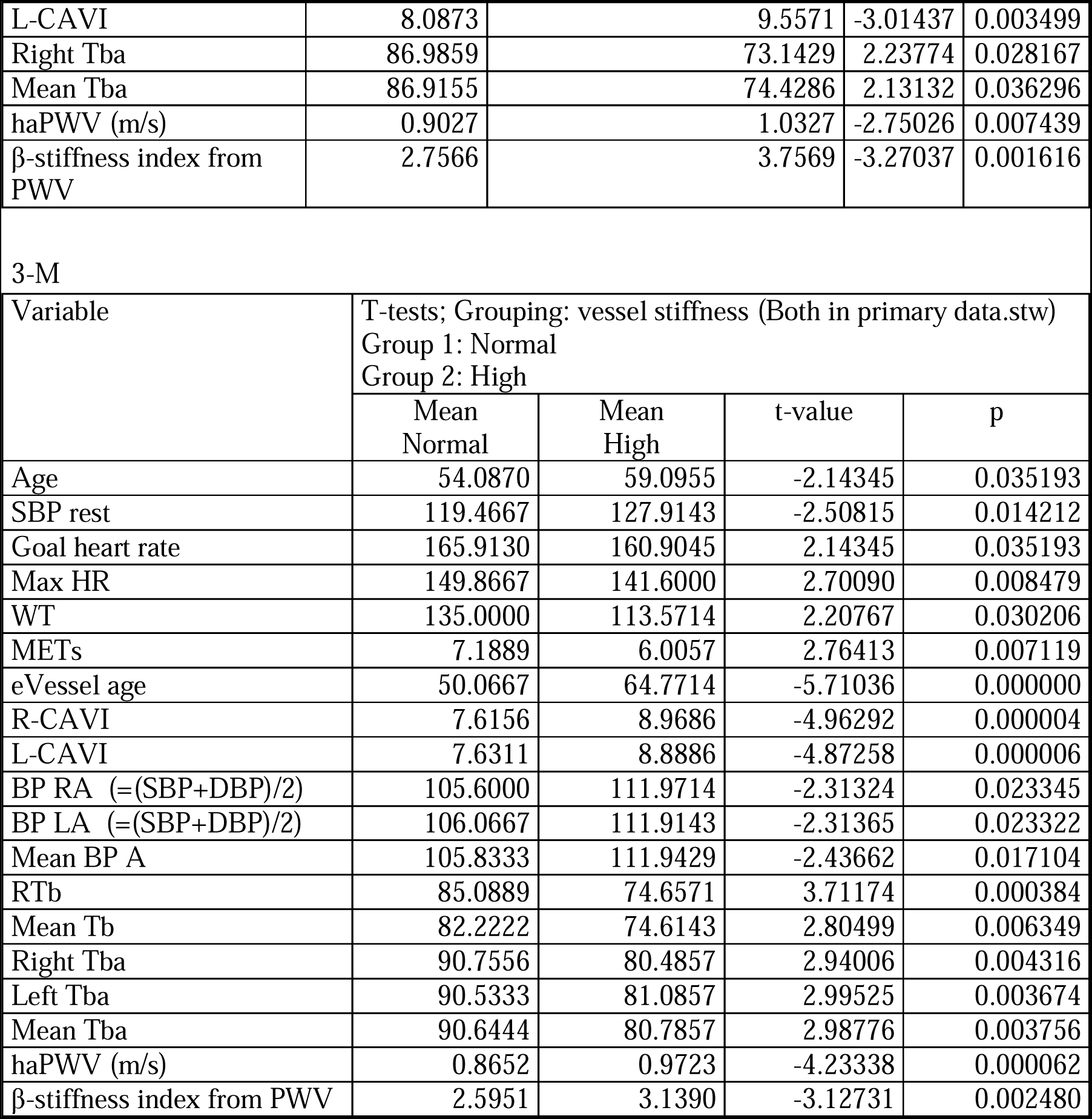
The comparative features of the sample divided by various binary categorical variables. The represented continuous variables are all statistically significant at p<0.05.

Patients with atherosclerosis of coronary artery have a higher CAVI in compare to patients without atherosclerosis of the coronary artery disease, R-CAVI; 9.0419 vs 7.6796, L-CAVI; 7.7122 vs 8.9226, p <0.000005, 0.000021, respectively.

Hemodynamically significant coronary artery atherosclerosis (>60%) exist in group with stress induced myocardial perfusion defect on the CTP imaging with ATP.

In terms of the correlations, the whole correlations between the continuous variables represented in the supplementary files. (*Suppl. file 1,2*)

The regression analysis showed no effect of the CAVI on the presence or absence of the IHD. (*Table 4*)

**Table 4:**
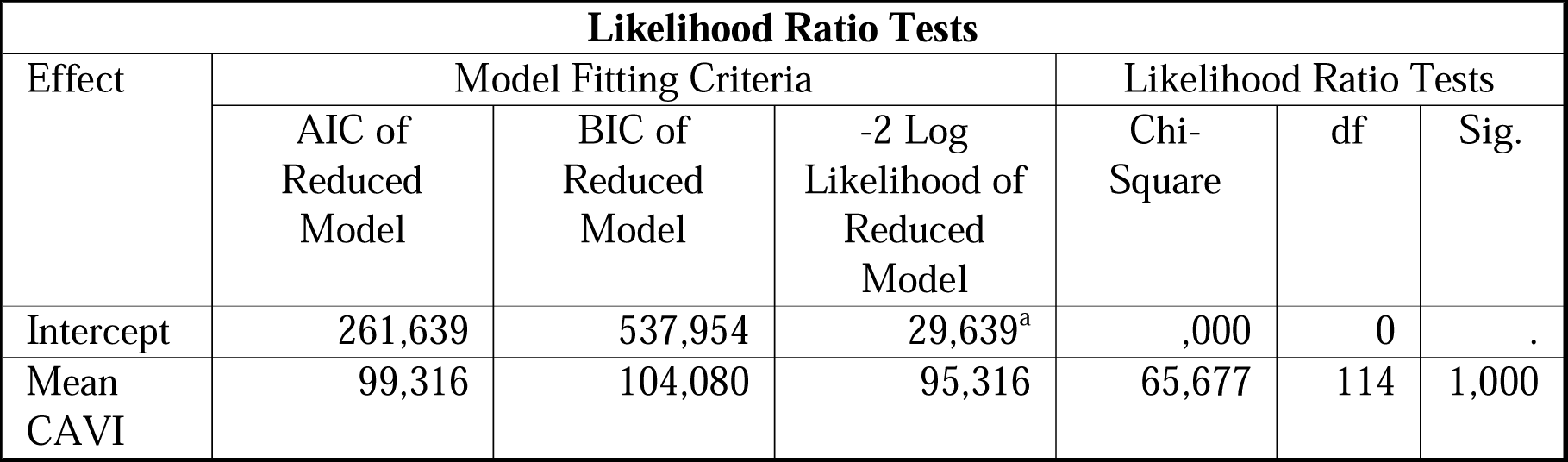
The chi-square statistic is the difference in -2 log-likelihoods between the final model and a reduced model. The reduced model is formed by omitting an effect from the final model. The null hypothesis is that all parameters of that effect are 0. a. This reduced model is equivalent to the final model because omitting the effect does not increase the degrees of freedom.

Moreover, the ROC analysis showed a 64 % diagnostic accuracy for IHD using CAVI parameter. (*Figure 1*)

**Figure 1:**
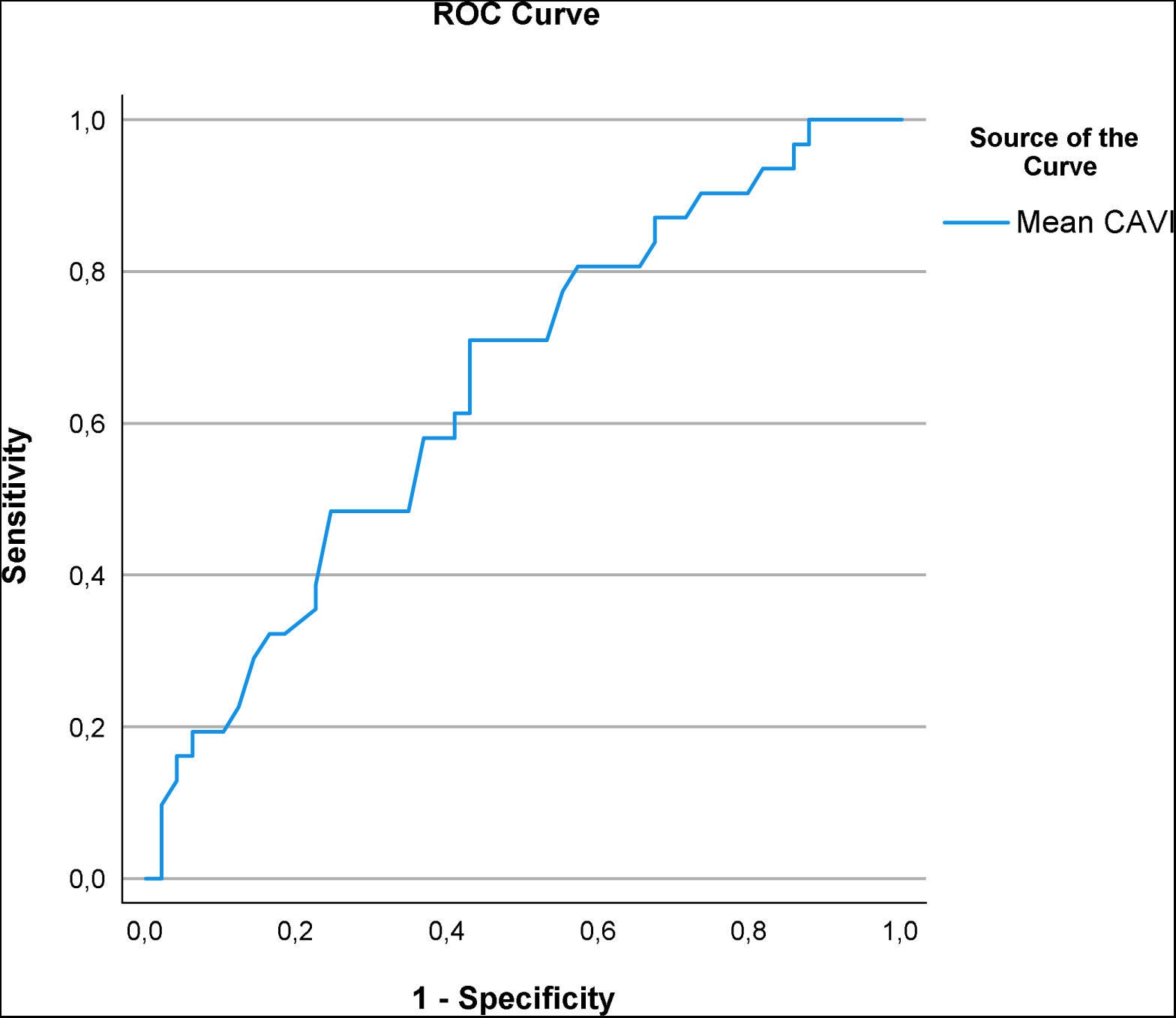
The CAVI parameter showed a 64 % Area Under the ROC Curve. The test result variable(s): Mean CAVI has at least one tie between the positive actual state group and the negative actual state group.

## The CAVI parameters assessment and features

Participants passed vessel stiffness test and pulse wave as well as vascular age by using Fukuda Denshi device (VaSera VS-1500; Japan). Cuffs placed to assess the vascular stiffness and the vascular age as well as the ancle-brachial index.

Cuffs fitted to the size of the arms and ankles of the patients. Electrodes attached to the two arms, and a microphone for cardio-phonogram measurements fixed with double-sided tape over the sternum in the second intercostal space. Cardio-ankle vascular index (CAVI) reflects the overall stiffness of the aorta, femoral artery and tibial artery, and is theoretically not affected by blood pressure [6]. CAVI measurements considered valid only when obtained during at least three consecutive heartbeats [6].

Four cuffs are positioned on the two arms, near the elbow flexion crease, and on the two legs, close to the ankles. This device performs all functions automatically, including the cuffs inflating and recording pulse waves with an incorporated oscillometric sensor. Using the VaSera VS-1500, we assessed the estimated vessel age, vessel stiffness right and left CAVI, right and left ABI, mean brachial SBP/ DBP, mean right and left brachial BP (BP RB =(RBSBP+LBDBP)/2); (BP LB =(LBSBP+LBDBP)/2), mean right and left ankle BP (BP RA =(RASBP+RADBP)/2); (BP LA =(LASBP+LADBP)/2), mean ankle BP, Mean ABI, right and left brachial pulse (RTb, LTb), mean brachial pulse (Tb), right and left brachial-ankle pulse (RTba, LTba), mean Tba. Then we calculated the length heart-ankle (Lha), heart-ankle pulse wave velocity (haPWV; m/s), and β-stiffness index from PWV.

The heart ankle pulse wave velocity (haPWV; m/s) is estimated using the following equation: heart-ankle length (Lha)/time. Time is the pulse transit time of the pulse wave from the origin of the aorta (peak of the R wave in lead II of an electrocardiographic record incorporated in the system) to its arrival at each of the extremities (the cuff placement). The Lha calculated by applying the next mathematical height-based formula: Lhl=0.8129×height (cm)+12.328. This measurement method was validated by VOPITB in the Spanish population and is also validated by VaSera VS-1500 [12]. Because VOPITB can measure the haPWV in each limb, CAVI can be determined. CAVI and haβ reflect the arteriosclerosis of the aorta, femoral artery and tibial artery quantitatively [13].

To make the best use of the nature of the stiffness parameter β to be used in VaSera, fixing the coefficients or termination of it is use considered [14]. Therefore, we used the β-stiffness index from haPWV (haβ) using the formula; haβ =2* ρ *(haPWV)^2*LN(SBP / DBP)/(SBP-DBP) Where; ρ is blood density (fixed value of 1050 kg/m3 in VaSera devices), Ln is natural logarithm, SBP is systolic blood pressure (SBP), and DBP is diastolic blood pressure (DBP). 1 mmHg is converted to 133.32 Pa.

CAVI is disregarded if the ABI is[<0.9. The main principle of the Fukuda Denshi device is by calculating the cardio ankle vascular index (CAVI) which is preferred on the local pulse wave velocity (PWV) devices. Additionally, CAVI is independent from blood pressure or much less sensitive to the blood pressure values than PWV [7,15]. The CAVI is measured when used the Fukuda Denshi device. The CAVI is preferred in compare to other PWV by higher accuracy in the determination of the risk of the cardiovascular disease [16,17]. Using the Fukuda Denshi device (VaSera VS-1500), the pulse wave velocity can be measured using the following formula haPWV=Lha/(Ta+Tba or Tb+Tba) Where; Tb; Brachial pulse Tba; tibial pules at brachial ankle Lha; heart ankle length Other CAVI measurements performed using VaSera VS-1500 according to the manufacturer’s instructions [13].

According to the Japanese Society for Vascular Failure, the normal CAVI index considered < 8, borderline: 9 > CAVI ≥ 8 from, pathological: CAVI ≥ 9. Normal ankle brachial index ABI considered from 0,9-1,29 [18]. According to the American heart association (AHA), ankle brachial index (ABI) readings are categorized as abnormal (ABI ≤0.90), borderline (ABI 0.91-0.99), and normal (ABI 1.00-1.40). An ABI >1.40 is considered noncompressible [18].

## Discussion

Using physical stress tested monitored 12 lead-ECG remains the elementary test for the primary detection of ischemic heart disease. However, severe limitations exist in the diagnostic accuracy related to the ECG artifact during the movement of the patients during the physical stress test.

Improving the diagnostic accuracy of the physical stress test is a point of focus of the cardiological scientific community. Several attempts performed to enhance the diagnosis performance of the physical exertion tests using complementary methods such as the dynamics of cardiac electrical activity (EAS) during exercise testing [19]. The study suggests that incorporating the equivalent electric cardiac generator of dipole type during exercise ECG testing can enhance the accuracy of diagnosing coronary artery disease [19].

Additional studies suggested the use of the single channel electrocardiography and or the exhaled volatile organic compounds as a molecular biomarker to detect the IHD [20–23].

The current study tried to demonstrate the strengthen and the benefits of the CAVI parameter in the diagnosis of IHD and or risk scoring of the IHD developing in the upcoming near future. Several studies demonstrated that CAVI can be a predictable for the cardiovascular diseases including the IHD [24]. Our study demonstrated a good diagnostic accuracy for the IHD (64 %) in compare to the physical stress test (55%), which is used as a classical method for IHD early detection [20]. The CAVI test is less time-consuming procedure in compare to the bicycle ergometry.

Single study demonstrated that high CAVI is associated with the higher post coronary artery bypass grafting surgery mortality [25].

With aging a dramatical elevation in the arterial stiffness and pulse pressure which is associated with increase of the systolic blood pressure and reduction of the diastolic blood pressure independence of the health and risk factors of the cardiovascular system. Our study showed similar findings that aging is associated with increase the CAVI levels and the further associated cardiovascular events.

Patients with a CAVI value[≥[10 have high incidence of heart disease and cerebrovascular events during the 3 year follow-up period with a cumulative incidence rate > 17% [26]. Therefore, CAVI is recommended to be applied in primary and secondary prevention in patients with cardiovascular risk factors and/or diseases to detect sub-clinical arterial alterations and to evaluate the effect of treatment and its monitoring [7].

Patients with stable coronary artery disease have a low eGFR as an indicator of IHD associated systemic changes including the changes in the kidney filtration function. Previous studies demonstrated the role of the myocardial infraction in the development of kidney dysfunction in terms of elevation of creatinine plasma level [27].

Several ongoing clinical trials to assess the reliability of single channel ECG in the diagnosis of ischemic heart disease and arrythmia in both adults and children (NCT05756309, NCT06181799).

## Conclusions

The CAVI showed no statistically significant difference between the two groups. According to the results of the multivariate regression analysis, CAVI levels not associated with the stress induced myocardial perfusion defect on the CTP. Therefore, CAVI levels are not potential tool to assess the risk of IHD development. Moreover, no correlation between the CAVI index and the stress induced myocardial perfusion defect on the CTP. However, the ankle-brachial index showed statistically significant difference between the patients with IHD and patients without IHD. Moreover, the estimated arterial age in the IHD group is statistically significantly higher than in the individuals without IHD. The CAVI shoed 64 % diagnostic accuracy for the stress induced myocardial perfusion defect on the CTP. Using the other possibilities of the Fukuda Denshi device parameters that can be calculated or estimated, additional indicators can be associated with the IHD include the Tba and haPWV parameters, which are higher in the IHD patients in compare to the non IHD group. Further, study on a larger sample is ongoing on clinicaltrials.gov (NCT06181799).

## Supporting information

Suppl. file 2. correlation in figures

Suppl. file 1. correlation map

## Data Availability

All data produced in the present work are contained in the manuscript

## List of abbreviations

CVD: cardiovascular disease
CTP: stress computed tomography myocardial perfusion imaging
IHD: ischemic heart disease

## Decelerations

### 1. Ethics approval and consent to participate

the study approved by the Sechenov University, Russia, from “Ethics Committee Requirement № 19-23 from 26.10.2023”. A written consent is taken from the study participants

### 2. Consent for publication

applicable on reasonable request

### 3. Availability of data and materials

applicable on reasonable request

### 5. Funding’s

The work of Philipp Kopylov and Daria Gognieva was financed by the government assignment 1023022600020-6 «Application of mass spectrometry and exhaled air emission spectrometry for cardiovascular risk stratification». The work of Basheer Marzoog and Peter Chomakhidze was financed by the Ministry of Science and Higher Education of the Russian Federation within the framework of state support for the creation and development of World-Class Research Center ‘Digital biodesign and personalized healthcare’ № 075-15-2022-304.

### 6. Authors’ contributions

MB is the writer, researcher, collected and analyzed data, interpreted the results, and revised the final version of the manuscript, PCh revised the paper, and PhK revised the final version of the manuscript. All authors have read and approved the manuscript.

## 7. Acknowledgments

not applicable

## 8. Authors’ information

**Basheer Abdullah Marzoog**, World-Class Research Center «Digital Biodesign and Personalized Healthcare», I.M. Sechenov First Moscow State Medical University (Sechenov University), 119991 Moscow, Russia; postal address: Russia, Moscow, 8-2 Trubetskaya street, 119991. (, +79969602820). ORCID: 0000-0001-5507-2413. Scopus ID: 57486338800. **Daria Gognieva**, World-Class Research Center «Digital Biodesign and Personalized Healthcare», I.M. Sechenov First Moscow State Medical University (Sechenov University), 119991 Moscow, Russia; postal address: Russia, Moscow, 8-2 Trubetskaya street, 119991. ORCID: 0000-0002-0451-2009. **Peter Chomakhidze**, World-Class Research Center «Digital Biodesign and Personalized Healthcare», I.M. Sechenov First Moscow State Medical University (Sechenov University), 119991 Moscow, Russia; postal address: Russia, Moscow, 8-2 Trubetskaya street, 119991. ORCID: 0000-0003-1485-6072. **Philipp Kopylov,** director of the institute of the World class Research Center «Digital Biodesign and Personalized Healthcare», I.M. Sechenov First Moscow State Medical University (Sechenov University), 119991 Moscow, Russia; postal address: Russia, Moscow, 8-2 Trubetskaya street, 119991. ORCID: 0000-0002-4535-8685. Scopus ID: 6507736224.

9.The paper has not been submitted elsewhere

## STANDARDS OF REPORTING

STROBE guideline has been followed.

