## Supplementary material for "Cardi-Ankle Vascular Index Optimizes Ischemic Heart disease Diagnosis": Suppl. file 2. correlation in figures

The statistically significant correlation between the CAVI and the other continuous parameters represented in the current supplementary file.

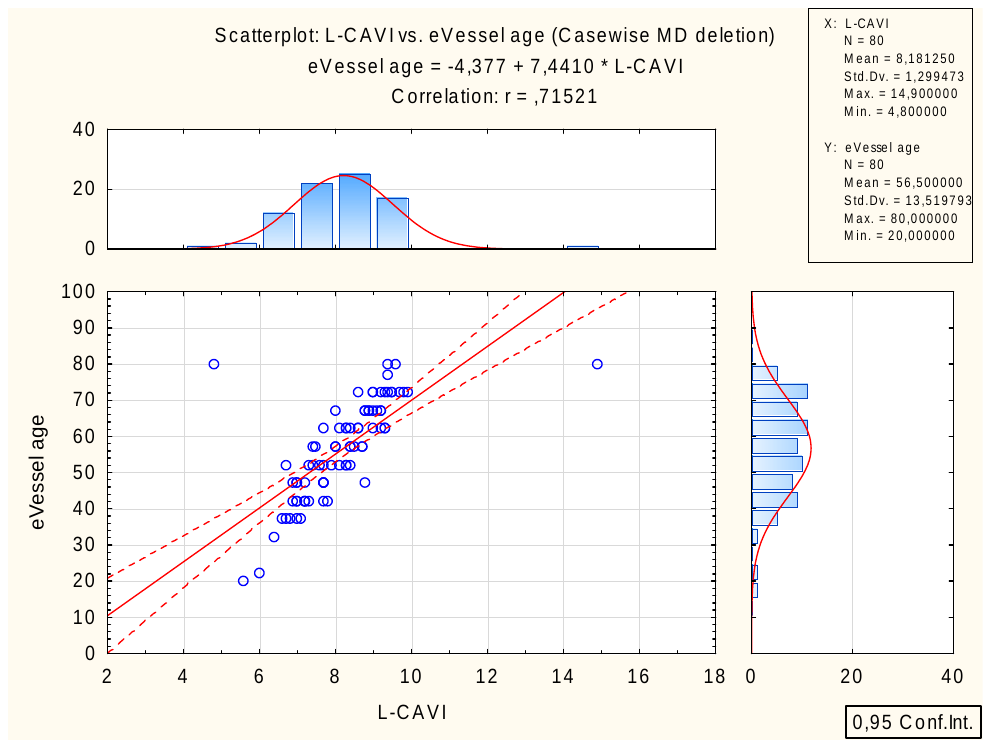

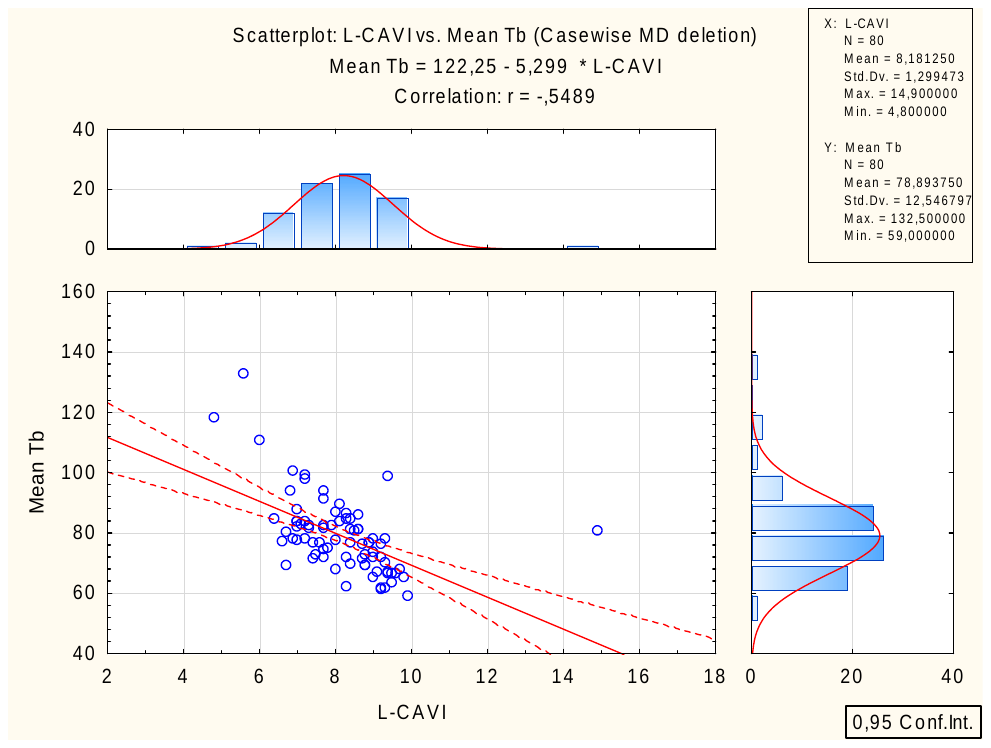

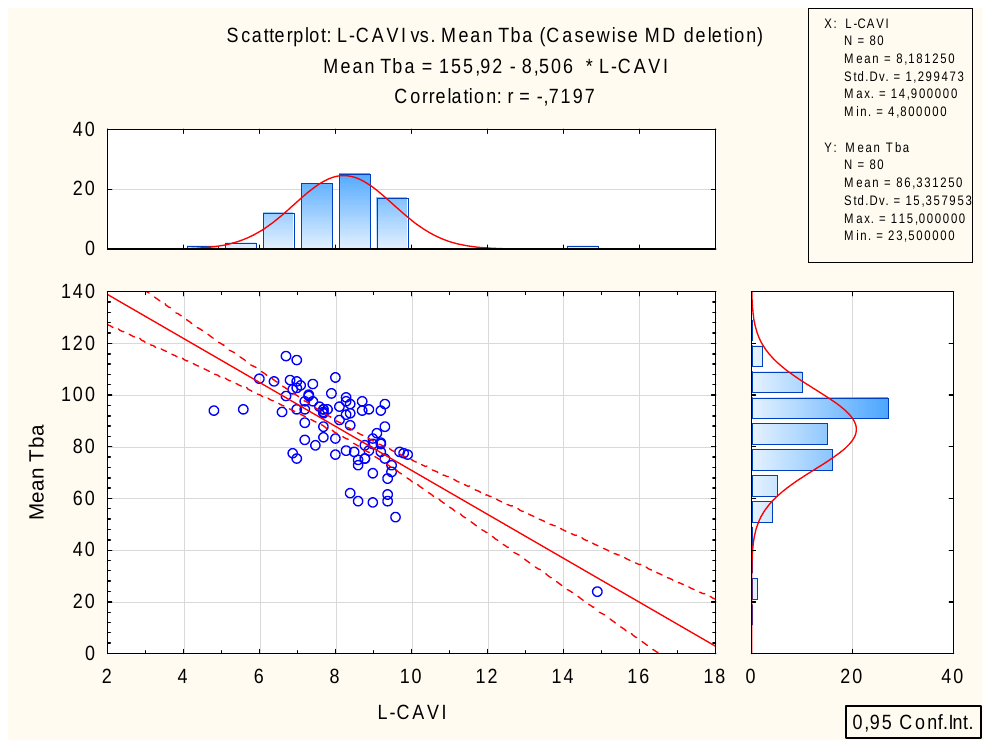

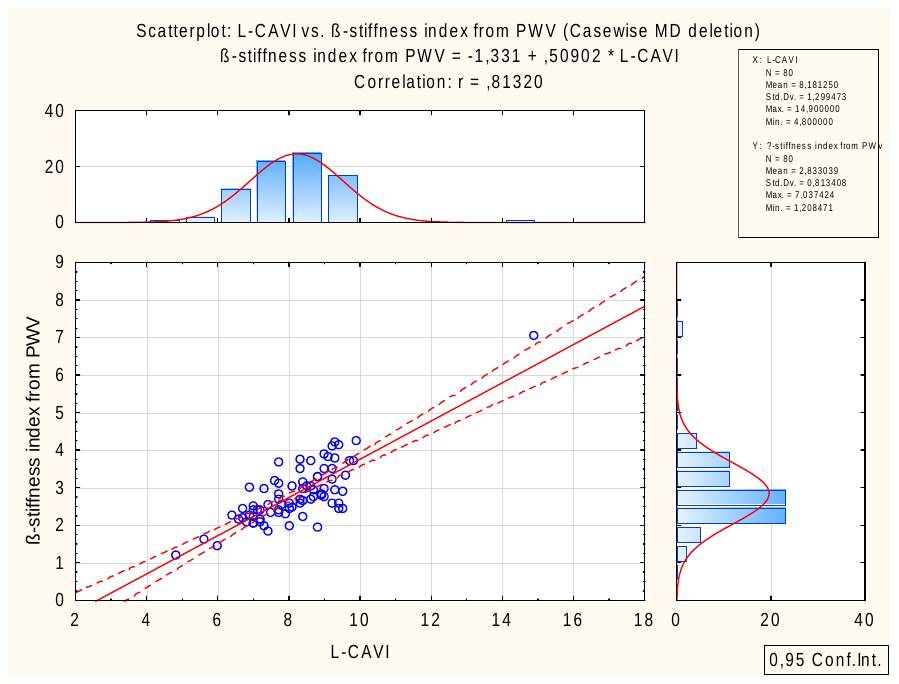

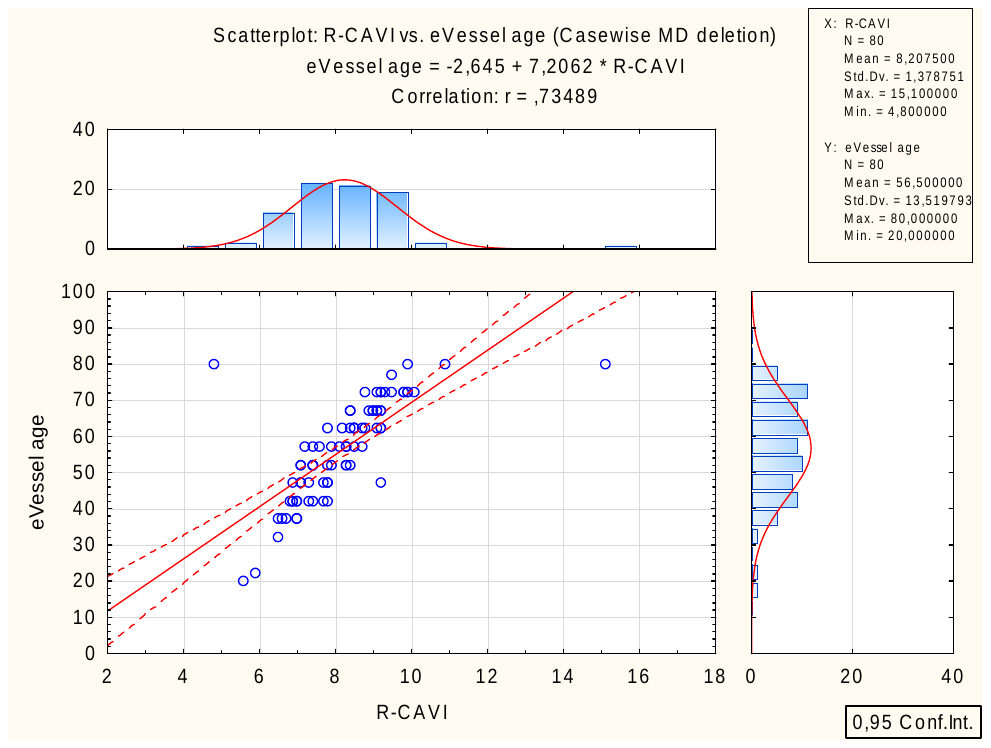

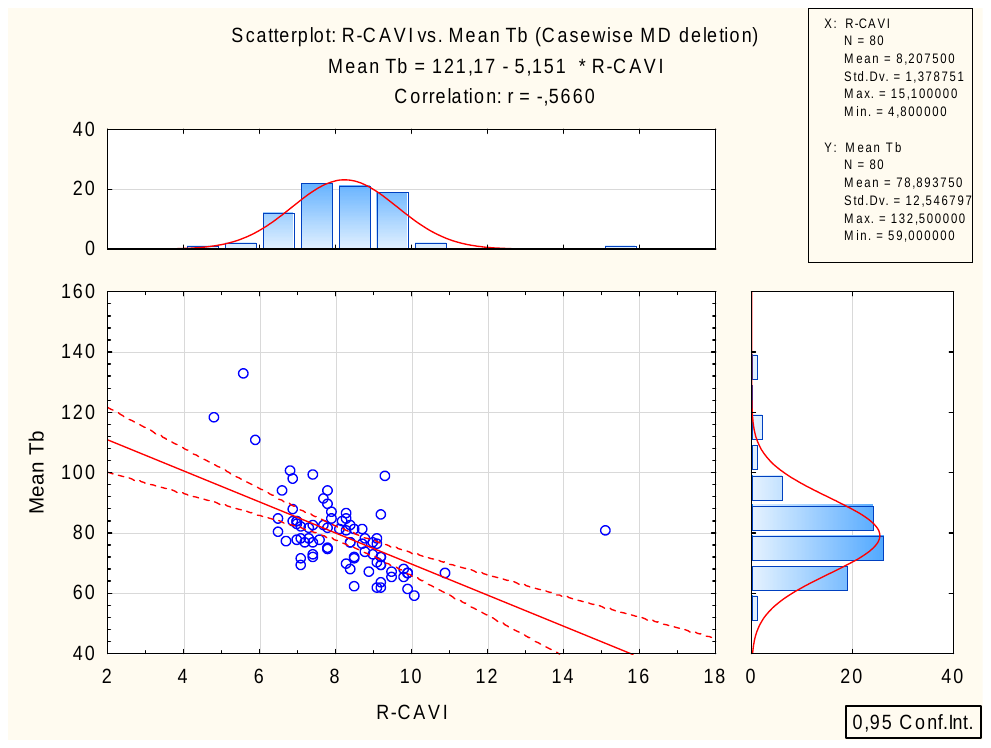

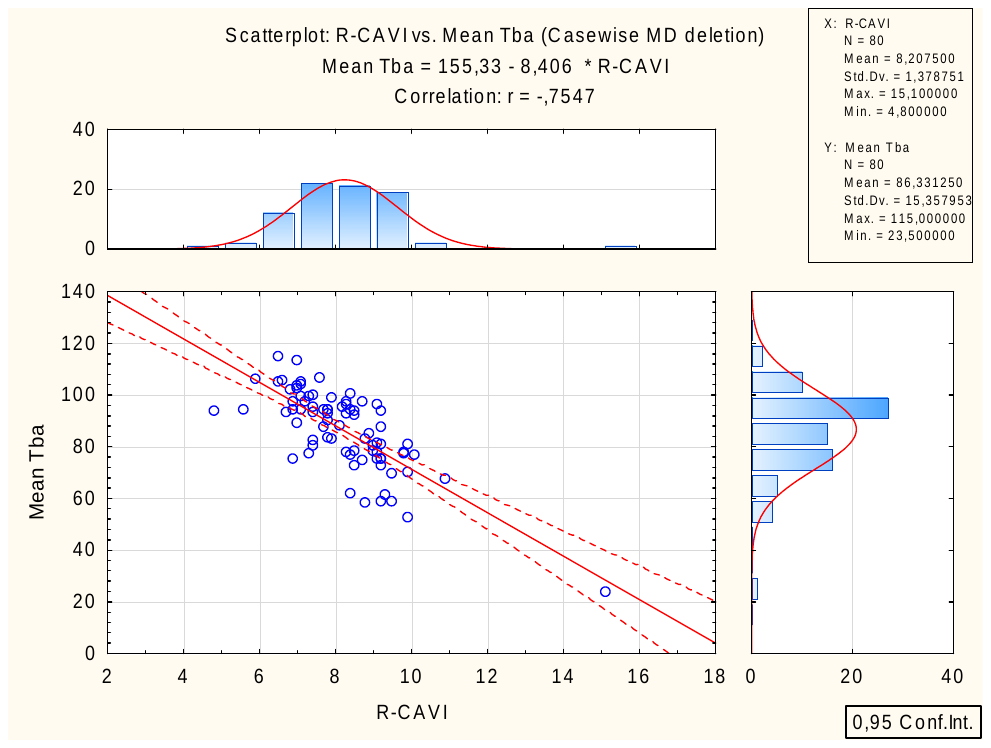

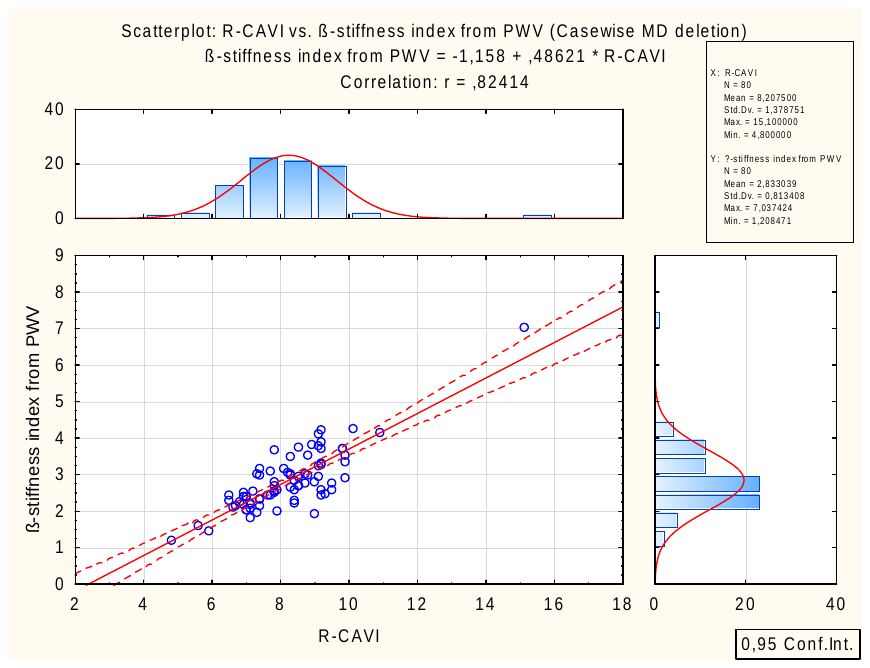

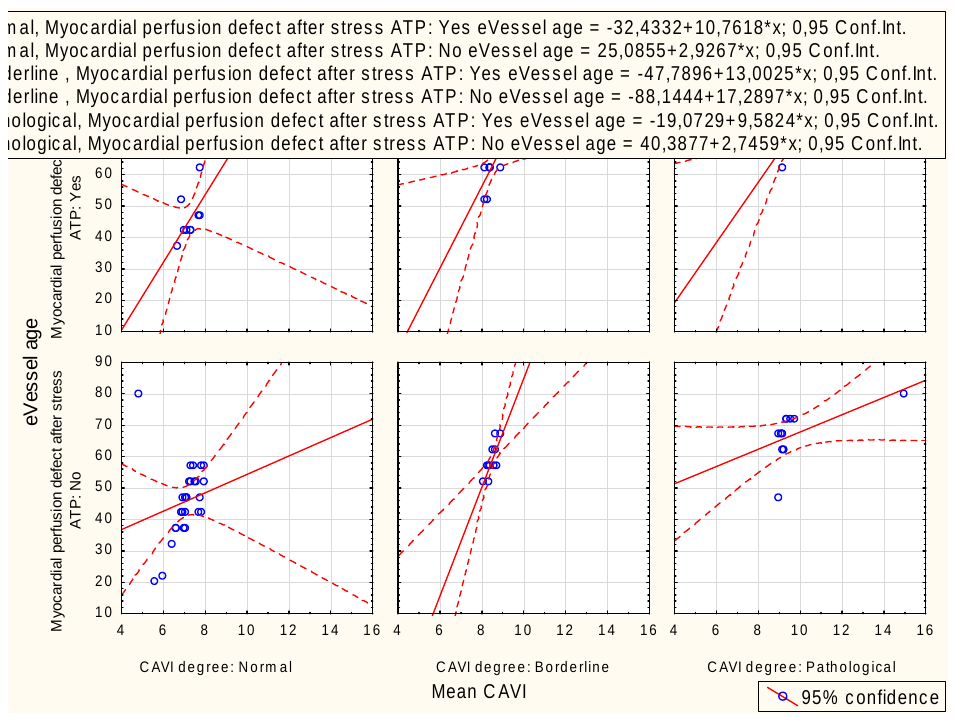

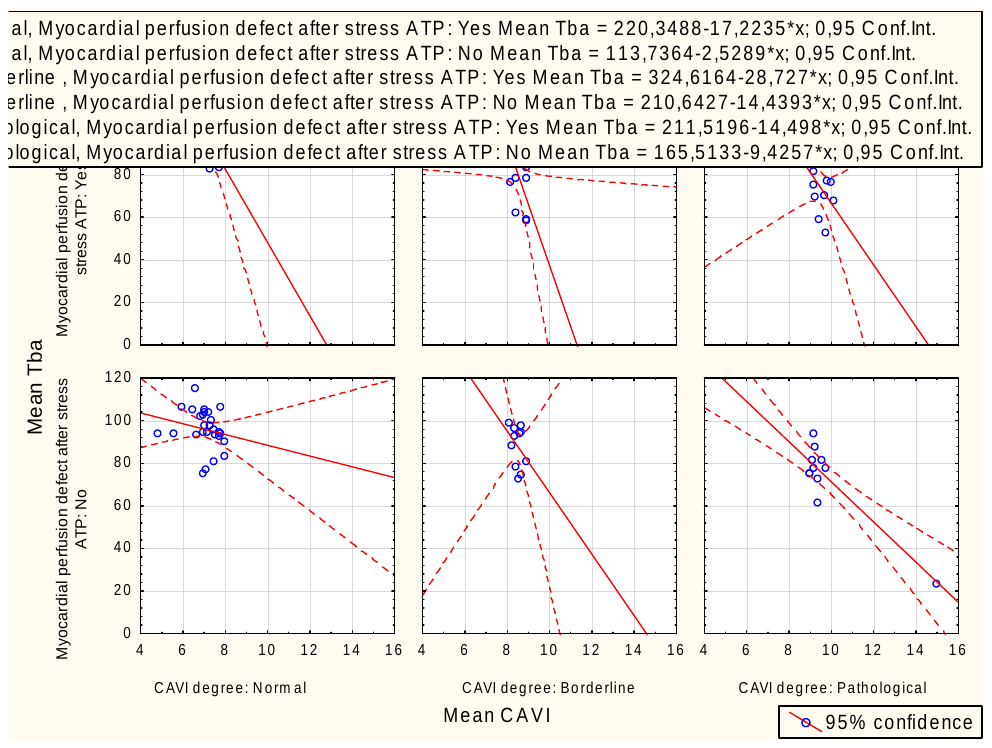

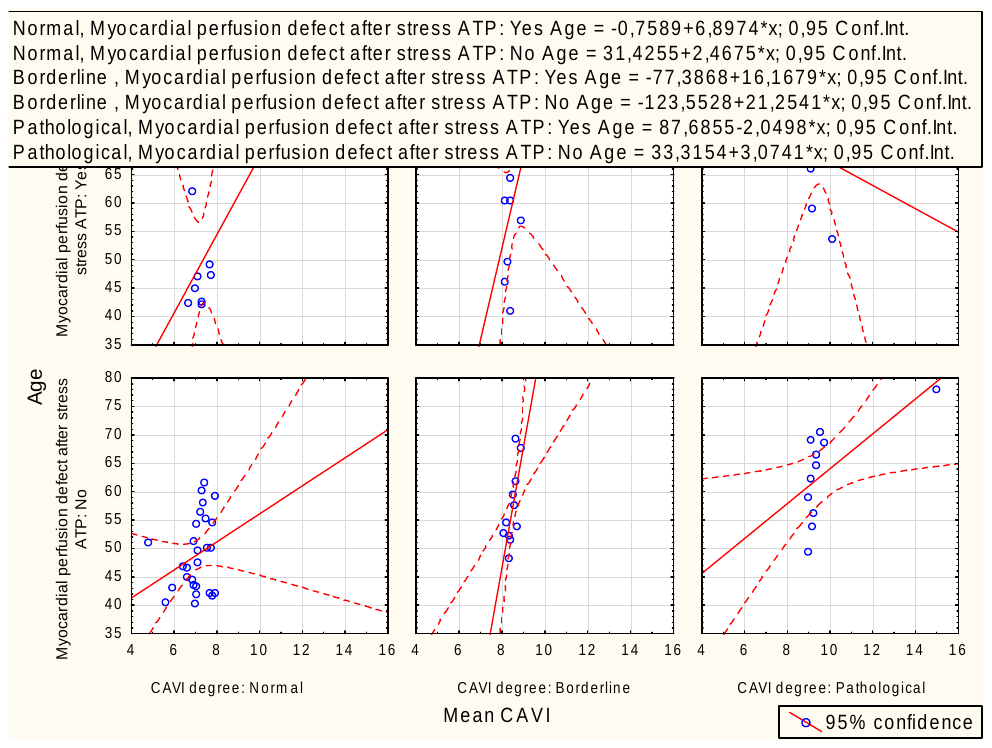
